# ME/CFS and PASC Patient-Derived Immunoglobulin Complexes Disrupt Mitochondrial Function and Alter Inflammatory Marker Secretion

**DOI:** 10.1101/2025.08.06.25332978

**Authors:** Zheng Liu, Claudia Hollmann, Sharada Kalanidhi, Stephanie Lamer, Andreas Schlosser, Emils Edgars Basens, Georgy Nikolayshvili, Liba Sokolovska, Gabriela Riemekasten, Rebekka Rust, Judith Bellmann-Strobel, Friedemann Paul, Robert K Naviaux, Zaiga Nora-Krukle, Franziska Sotzny, Carmen Scheibenbogen, Bhupesh K Prusty

## Abstract

Autoimmunity is a key clinical feature in both post-infectious Myalgic encephalomyelitis / chronic fatigue syndrome (ME/CFS) and Post-Acute Sequelae of COVID (PASC). Passive transfer of immunoglobulins from patients’ sera into mice induces some clinical features of PASC. IgG-induced transfer of disease phenotypes has long been appreciated, yet the exact mechanism of disease development remains largely elusive. Here, we demonstrate that IgG isolated from post-infectious ME/CFS patients selectively induces mitochondrial fragmentation in human endothelial cells, thereby altering mitochondrial energetics. This effect is lost upon cleavage of IgG into its Fab and Fc fragments. The digested Fab fragment from ME/CFS alone was able to alter the mitochondrial energetics, resembling the effect of intact IgG. In contrast, the Fc fragment alone induced a hypometabolic phenotype characterized by a trend towards reduced overall ATP content. IgG from ME/CFS and PASC patients induced distinct but separate cytokine secretion profiles in healthy PBMCs. Proteomics analysis of IgG-bound immune complexes revealed significant changes within the immune complexes of ME/CFS patients, affecting extracellular matrix organization, while the same from PASC patients pointed towards alterations in hemostasis and blood clot regulation. We demonstrate that IgGs from ME/CFS patients carry a chronic protective stress response that promotes mitochondrial adaptation via fragmentation, without altering mitochondrial ATP generation capacity in endothelial cells. Together, these results highlight a potential pathogenic role of IgG in post-infectious ME/CFS and point to novel therapeutic strategies targeting antibody-mediated metabolic dysregulation.

**One Sentence Summary:** IgG immune complexes from ME/CFS and PASC patients differ from those of healthy individuals and affect mitochondrial structure and function.

## INTRODUCTION

Myalgic Encephalomyelitis/Chronic Fatigue Syndrome (ME/CFS) is a complex, debilitating, chronic, long-term illness that involves immune dysfunction, autonomic nervous system dysfunction, and metabolic alterations (*1-3*). Key symptoms of ME/CFS include fatigue, which can be both mental and physical in nature, such as post-exertional malaise (PEM) (*4*), cognitive dysfunction (“brain fog”), etc (*5*). Mental fatigue can originate from alterations in specific regions of the brain that affect the function of neurotransmitters (*6*), while physical fatigue can result from metabolic changes within muscle cells (*7*).

Mitochondria play a crucial role in energy production by generating ATP through the process of oxidative phosphorylation. Mitochondrial dysfunction can lead to impaired ATP production, increased oxidative stress, and metabolic dysregulation (*8, 9*). Increasing evidence suggests that abnormal mitochondria play a crucial role in the development and/or progression of ME/CFS. Alterations in mitochondrial dynamics, deficient mitochondrial ATP generation, altered mitochondrial and cellular bioenergetics, and increased oxidative stress during ME/CFS have been reported (*2, 10-16*). Biophysical changes in cells from ME/CFS patients placed under osmotic stress have recently been used as an innovative diagnostic test for ME/CFS (*17*). Major alterations in the metabolic profile of ME/CFS patients have been reported. Such alterations can be directly associated with changes in mitochondrial and glycolytic metabolic pathways (*18, 19*). Key symptoms of ME/CFS could also be explained by mitochondrial dysfunction. In muscle cells, mitochondrial dysfunction leads to disorders including mitochondrial myopathy, Duchenne muscular dystrophy, cachexia, and sarcopenia. These conditions are characterized by reduced ATP production and increased oxidative stress, leading to muscle weakness, fatigue, and degeneration (*20-22*). In neural cells, ATP production, calcium homeostasis, membrane excitability, and neurotransmission regulation are also highly dependent on mitochondria (*23, 24*). Taken together, mitochondrial dysfunction could contribute to both physical and neurological issues in ME/CFS. However, the exact antecedent to mitochondrial modulation in ME/CFS is largely unknown.

ME/CFS patients frequently show altered regulation of the autonomic nervous and immune system (*25*). In recent years, several studies have focused on the role of autoimmunity in ME/CFS, showing elevated levels of antibodies targeting β2 adrenergic receptors (β2 AdR) and muscarinic M3 and M4 acetylcholine receptors in subsets of ME/CFS patients (*26-29*). The correlation of these autoantibodies with clinical symptoms suggests their functional role in ME/CFS (*30*). Two recent studies reported the induction of ME/CFS and long COVID-like symptoms in mice following the passive transfer of IgG from patients (*31, 32*). These findings collectively highlight the potential role of autoantibodies in the pathophysiology of ME/CFS, particularly in disrupting the regulation of autonomic and immune systems. In this study, we demonstrate the role of human IgG in mitochondrial alterations *in vitro*.

## RESULTS

### Passive transfer of IgGs from ME/CFS patients induces mitochondrial fragmentation

Immune dysregulation is closely associated with mitochondrial dysfunction, which is a characteristic feature of ME/CFS. Several groups, including ours, have previously shown that serum-derived unknown factors from ME/CFS patients can induce mitochondrial alterations in healthy cells (*10, 17, 33*). A lack of complement activation and pathogen clearance can allow several opportunistic pathogens to replicate, resulting in the production of a wide array of IgG and IgM autoantibodies in the serum. We hypothesized that some of the altered antibodies in serum or altered antigen-antibody complexes in might induce chronic mitochondrial dysfunction. To investigate this *in vitro*, we purified IgGs (Figure 1A) and exposed them to two independent cell types, primary human foreskin fibroblasts (HFFs) and primary human umbilical vein endothelial cells (HUVECs), and followed their entry into the cells using immunoblotting (Figure 1B). HUVECs were permissive to human IgG, while HFFs did not allow entry of IgG into the cells (Figure 1C), suggesting selective entry of IgG into human cells. IgG entry into HUVEC cells was also confirmed using immunofluorescence staining of human IgG (Figure 1D). IgG that entered HUVEC cells was efficiently detectable up to 16 h post-exposure (Figure 1E). However, IgG levels decreased inside the cells in a time-dependent manner (Figure 1E), suggesting potential recycling of IgG within the cell. Fc-specific fragments (∼35 kDa) of IgG remained abundantly detectable within the cells till 48 h post-exposure (Figure 1E). No major differences in IgG entry and survival of Fc-specific IgG fragments were observed between different disease groups (Figure 1F).

**Figure 1.**
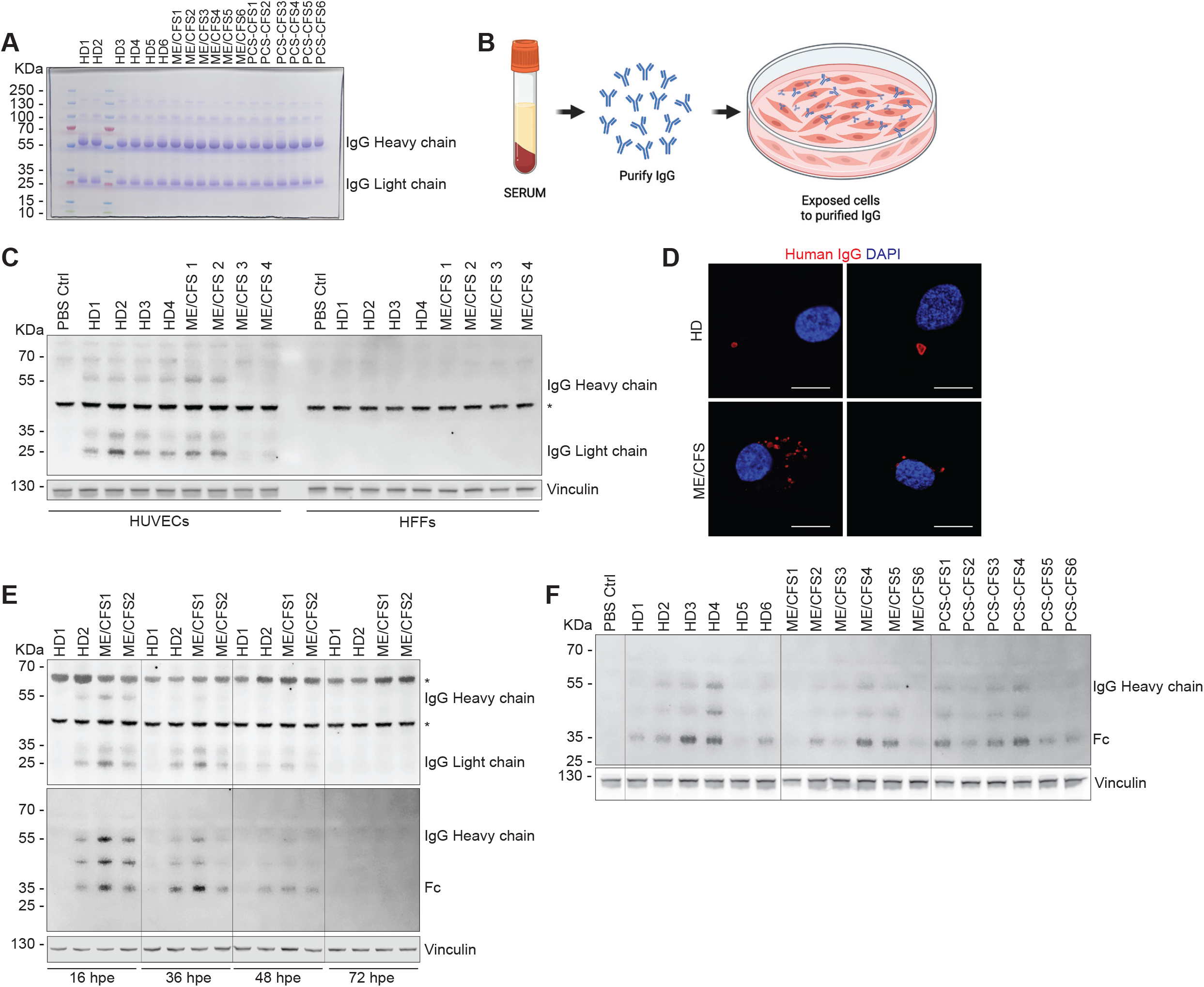
Selective entry of IgG into human cells and its turnover. A. The quality and quantity of purified IgG were assessed by running them on denaturing SDS-PAGE gels and staining with Coomassie. B. Schematics of IgG entry assay. C. Immunoblot analysis reveals the presence of intracellular IgG heavy and light chains in HUVECs, but not in HFFs, following exposure to 1 μg/mL purified IgG from ME/CFS patients or control sera for 12 hours. Vinculin was used as a loading control. D. Confocal microscopy images show intracellular human IgG in primary HUVECs exposed to 1⍰μg/mL purified IgG from patient and control sera. Two representative images are shown for each condition. E. Immunoblot analysis reveals degradation of intracellular IgG heavy and light chains in HUVECs at 16, 36, 48, and 72⍰h following exposure to 1⍰μg/mL purified IgG from ME/CFS patients or control sera. Vinculin was used as a loading control. Fc-specific secondary antibody was used (middle panel) to detect the full-length heavy chain of IgG and cleaved Fc fragments. F. Immunoblot analysis comparing intracellular IgG heavy and Fc-specific fragments in HUVECs at 16⍰h following exposure to 1⍰μg/mL purified IgG from HD, ME/CFS, and PCS-CFS patient sera. Vinculin was used as a loading control. Fc-specific secondary antibody was used to detect the full-length heavy chain of IgG and cleaved Fc fragments.

Exposure to as low as 1 µg/ml of IgGs from ME/CFS and PCS-CFS patients significantly fragmented mitochondria in HUVECs within 16 h of exposure (Figure 2B). Although the statistical significance was strong for the entire ME/CFS cohort (HD vs. ME/CFS, P=0.0004), IgG-induced mitochondrial fragmentation was not detected in the whole ME/CFS cohort. Only a subgroup of patients induced strong mitochondrial fragmentation. Hence, we conducted a gender-based analysis of the same, which revealed strong IgG-induced mitochondrial fragmentation in female ME/CFS and PCS-CFS patients compared to male patients (Figure 2C). IgG-induced mitochondrial fragmentation was also not observed when cells were exposed to IgGs from healthy controls and multiple sclerosis (Figures 2B, 2C). IgG from ME/CFS and PCS-CFS patients decreased the protein levels of Drp1, mitofilin, Miga1, and PLD6 (Figures 2D, 2E). Interestingly, the levels of the mitophagy marker protein LC3β showed a trend of decrease in the presence of IgG from ME/CFS and PCS-CSF patients (Figures 2D, 2E), but this difference was not statistically significant. Blocking the Fc receptors on HUVEC cell surface did not completely prevent IgG entry into cells (Figure 2F). Higher concentrations of Fc blocker showed decreased IgG entry (Figure 2F). However, there was significant cell loss at that concentration, as evident from a decrease in Drp1, p53, Tom20, and Vinculin protein levels. To understand the mechanism of IgG entry into cells, we digested IgG and separated both Fab and Fc fragments (Figure 2G). Both fragments were able to enter the cell with equal efficiency (Figure 2H). However, separated Fab and Fc fragments did not induce a fragmented mitochondrial phenotype (Figure 2I) with the same efficiency as their IgG counterpart. These results suggest that intact IgG from ME/CFS patients induces mitochondrial fragmentation, which might be independent of the classical Drp1-induced pathways.

**Figure 2.**
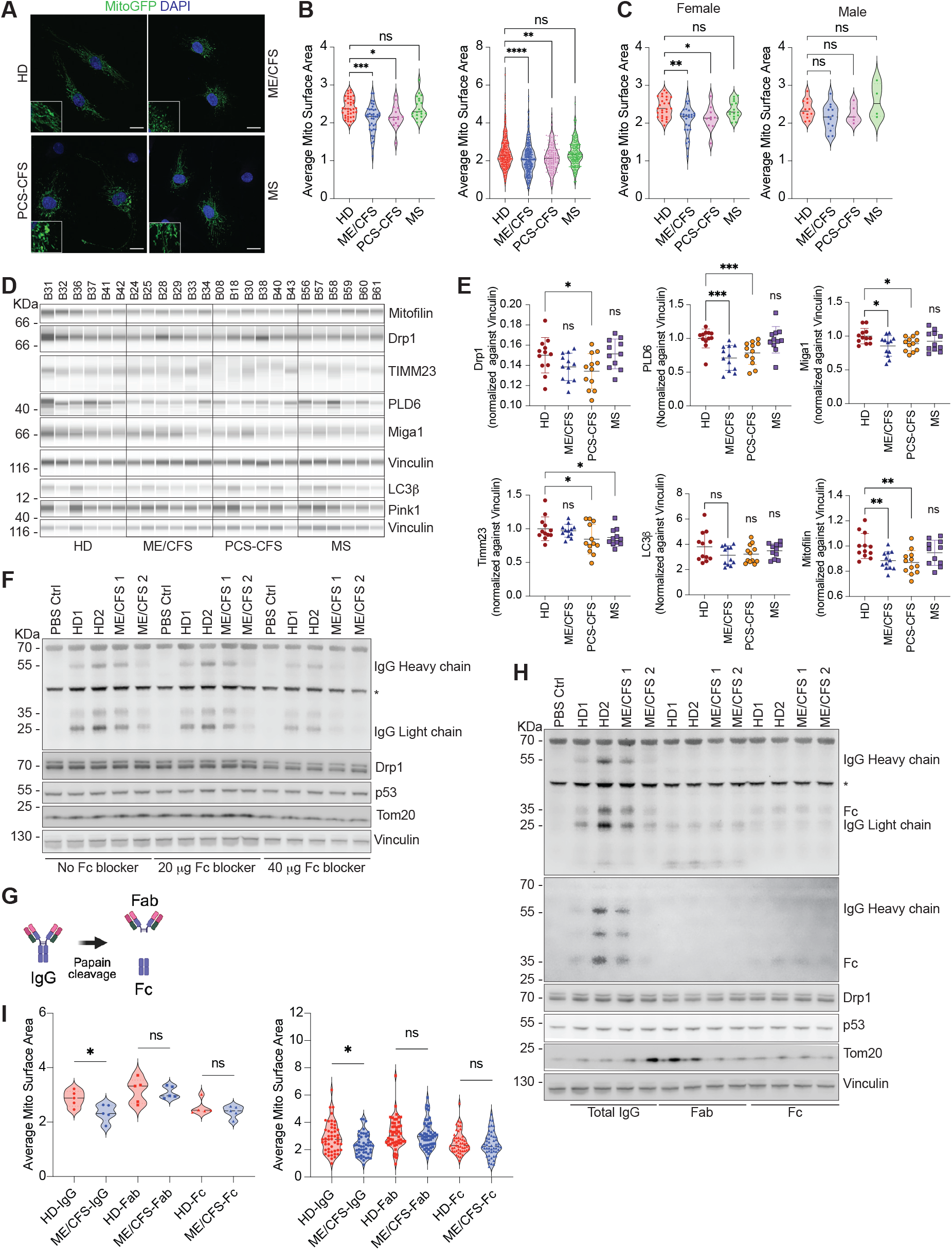
IgG from ME/CFS patients induces mitochondrial fragmentation. A. Representative confocal microscopy images show mitochondrial architecture in primary HUVECs expressing stable GFP within mitochondria and exposed to 1⍰μg/mL purified IgG from ME/CFS, PCS-CFS, and MS patient and control sera. One representative image is shown for each condition. B. Quantification of average mitochondrial surface area in primary HUVECs exposed to 1⍰μg/mL purified IgG from ME/CFS (n=39), PCS-CFS (n=15), MS (n=20) patients, and healthy controls (n=41). At least 10 cells were imaged for each sample derived from 3 replicates. In the left panel, each point represents the mean surface area across all images for each subject. In the right panel, each point represents the surface area measured from a single image. Two-tailed Mann-Whitney U test. (Left) HD vs ME/CFS, ***P=0.0004. HD vs PCS-CFS, *P=0164. HD vs MS, ns P=0.8676. (Right) HD vs ME/CFS, ****P<0.0001. HD vs PCS-CFS, **P=0.0067. HD vs MS, ns P=0.4794. C. Gender-based separation of average mitochondrial surface area from the above experiment. Each point represents the surface area measured from a single image. Two-tailed Mann-Whitney U test. (Female) HD vs ME/CFS, **P=0.0018. HD vs PCS-CFS, *P=0173. HD vs MS, ns P=0.4938. (Male) HD vs ME/CFS, ns P=0.0976. HD vs PCS-CFS, ns P=0.5058. HD vs MS, ns P=0.4462. D. Capillary-based automated Immunoblot analysis of Mitofilin, Drp1, TIMM23, PLD6, Miga, LC3b, Pink1 protein levels in HUVEC cells 36 hours after exposure to 1μg/mL of purified IgG from ME/CFS (n=12), PCS-CFS (n=12), and MS (n=11) patient and control (n=12) sera. Vinculin was used as a loading control. E. Fold change values from the above immunoblot analysis were derived from densitometric analysis of bands, normalized to the same value for vinculin. Two-tailed Mann-Whitney U test. (Drp1) HD vs ME/CFS, ns P=0.0684. HD vs PCS-CFS, *P=0387. HD vs MS, ns P=0.7859. (PLD6) HD vs ME/CFS, ****P<0.0001. HD vs PCS-CFS, ***P=0.0007. HD vs MS, ns P=0.6075. (Timm23) HD vs ME/CFS, ns P=0.1402. HD vs PCS-CFS, *P=0.0317. HD vs MS, *P=0.0106. (Miga1) HD vs ME/CFS, *P=0.0284. HD vs PCS-CFS, *P=0.0449. HD vs MS, ns P=0.1896. (LC3b) HD vs ME/CFS, ns P=0.0684. HD vs PCS-CFS, ns P=0.1600. HD vs MS, ns P=0.6075. (Mitofilin) HD vs ME/CFS, **P=0.0056. HD vs PCS-CFS, **P=0.0029. HD vs MS, ns P=0.3164. F. Immunoblot analysis shows a decreased amount of intracellular IgG heavy and light chains in HUVECs pretreated with Fc blocker before the exposure to 1⍰μg/mL purified IgG from ME/CFS patients or control sera for 12⍰h. Vinculin was used as a loading control. G. Schematics of IgG cleavage assay. H. Immunoblot analysis shows the presence of intracellular IgG heavy and light chains in HUVECs after exposure to 1⍰μg/mL purified IgG, purified Fab fragments, and Fc fragments from ME/CFS patients or control sera for 12⍰h. Vinculin was used as a loading control. Fc-specific secondary antibody was used (second panel) to detect the full-length heavy chain of IgG and cleaved Fc fragments. I. Quantification of average mitochondrial surface area in primary HUVECs exposed to 1⍰μg/mL purified IgG or Fab Fragment or Fc fragment from 5 ME/CFS, and 5 controls. In the upper panel, each point represents the mean surface area across all images for each individual. In the lower panel, each point represents the surface area measured from a single image. Two-tailed Mann-Whitney U test. (Upper Panel) HD vs ME/CFS IgG, *P=0.0317. HD vs ME/CFS Fab, ns P=0.8413. HD vs ME/CFS Fc, ns P=0.6905. (Lower Panel) HD vs ME/CFS IgG, *P=0.0130. HD vs ME/CFS Fab, ns P=0.5572. HD vs ME/CFS Fc, ns P=0.3662.

### IgGs from ME/CFS patients alters mitochondrial energetics

We then asked if IgG-induced changes in mitochondrial architecture also alter mitochondrial function. To assess the functional changes in the cellular metabolic profile, we performed mitochondrial stress tests using XFe96 Extracellular Flux Analyzer. Following 36 hours IgG exposure, HUVECs were analyzed for changes in oxygen consumption rate (OCR) and extracellular acidification rate (ECAR). Sequential injections of oligomycin, FCCP, and rotenone/antimycin A allowed for assessment of mitochondrial respiration parameters. Due to the pronounced and well-documented interplate variability in the seahorse assay, which is both observed in our data and others (*34, 35*), we carried out our assays in two separate groups. Each group included IgG from ME/CFS patients, either with a strong mitochondrial fragmentation phenotype or without it. On every single assay plate, we tested 10 healthy controls, 10 ME/CFS patients, along with either 10 PCS-CFS patients or 10 MS patients. IgG from healthy controls, PCS-CFS and MS patients induced similar OCR patterns in HUVECs, while HUVECs exposed to IgG from ME/CFS patients with mitochondrial fragmentation phenotype exhibited a trend towards higher basal respiration (not statistically significant) and significantly increased maximal respiration (Figures 3A) as observed from significantly higher spared respiration capacity. However, this phenomenon was not detected with IgG from ME/CFS patients without a mitochondrial fragmentation phenotype (Figure 3B). This suggests that IgG-induced mitochondrial fragmentation has a direct association with increased maximal respiration. In the meantime, ECAR, which indicates glycolytic activity, showed no differences in patterns. Baseline ECAR levels were comparable among all four patient groups (Figures 3C, 3D). ATP measurement assays did not reveal any significant changes in glycolytic, mitochondrial, or total ATP levels in response to IgG treatment (Supplementary Figures S1A-S1F).

**Figure 3.**
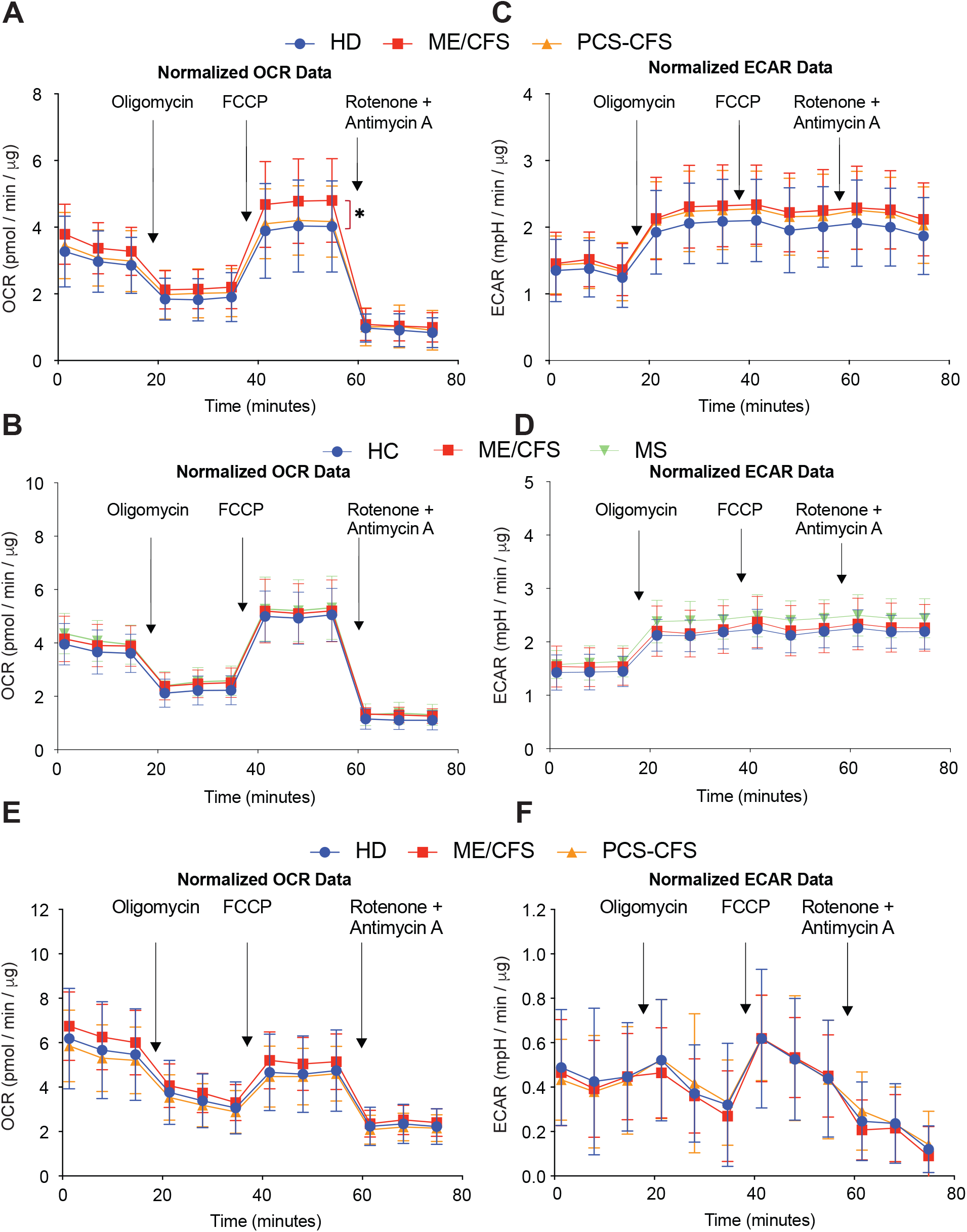
Patient IgGs alter metabolic energetics in HUVECs. A. A, B. Normalized OCR in HUVECs exposed to 1⍰μg/mL IgGs. C, D. Normalized ECAR in HUVECs exposed to 1⍰μg/mL IgGs. A-D. Seahorse assay mito stress test in the presence of medium containing glucose. ME/CFS (n=10), PCS-CFS patients (n=10), and healthy controls (n=10) were tested as three biological replicates for each subject. A, C. Representative ME/CFS patients are from the subgroup that induced strong mitochondrial fragmentation. B, D. Representative ME/CFS patients are from the subgroup that did not induce mitochondrial fragmentation. E. Normalized OCR in HUVECs exposed to 1⍰μg/mL IgGs. F. Normalized ECAR in HUVECs exposed to 1⍰μg/mL IgGs. E-F. Seahorse assay mito stress test in the presence of medium containing galactose. ME/CFS (n=10), PCS-CFS patients (n=10), and healthy controls (n=10) were tested as three biological replicates for each subject.

To force the cells to rely exclusively on oxidative phosphorylation (OXPHOS), we then replaced glucose-containing culture media with galactose-containing media, thereby isolating mitochondrial function without the influence of glycolysis. Due to the substitution with galactose, the cells had already reached their maximum respiratory capacity at baseline (Figure 3E). In contrast to the results from cells in glucose-containing media, HUVECs exposed to IgGs from the three study groups did not exhibit any differences in OCR or ECAR patterns (Figure 3F). These results showed that the ME/CFS IgG-induced increase in maximal respiratory capacity of cells is possibly not compensated by mitochondrial oxidative phosphorylation alone, and the observed effect is masked when cells are forced to rely solely on OXPHOS.

We then compared the potential effects of separated Fab and Fc fragments on mitochondrial energetics to those of the intact IgGs. While Fab fragments of IgG mimicked the intact IgG effects of ME/CFS patients (Figure 4A) as compared to healthy controls, the Fc fragments showed a completely opposite effect. Both the OCR and ECAR patterns of Fc-induced energetics remained low compared to those of healthy individuals, suggesting an overall decrease in mitochondrial function in the presence of ME/CFS Fc fragments, mimicking a hypometabolic phenotype. The glycolytic ATP and total ATP content in the presence of ME/CFS Fc fragment showed a trend towards hypometabolic phenotype (decreased ATP) (Figures 4B-4D) (HD vs. ME/CFS Fc, P=0.0952), but was not statistically significant. Taken together, our results suggest that IgG-induced alterations in mitochondrial morphology and energetics are potentially two independent phenotypes. While mitochondrial fragmentation was dependent on intact IgG, the effects of IgG and its various components (Fab and Fc) on cellular energetics varied, suggesting a complex relationship between circulating antibodies, associated immune complexes, and mitochondrial dysfunction and metabolic stress.

**Figure 4.**
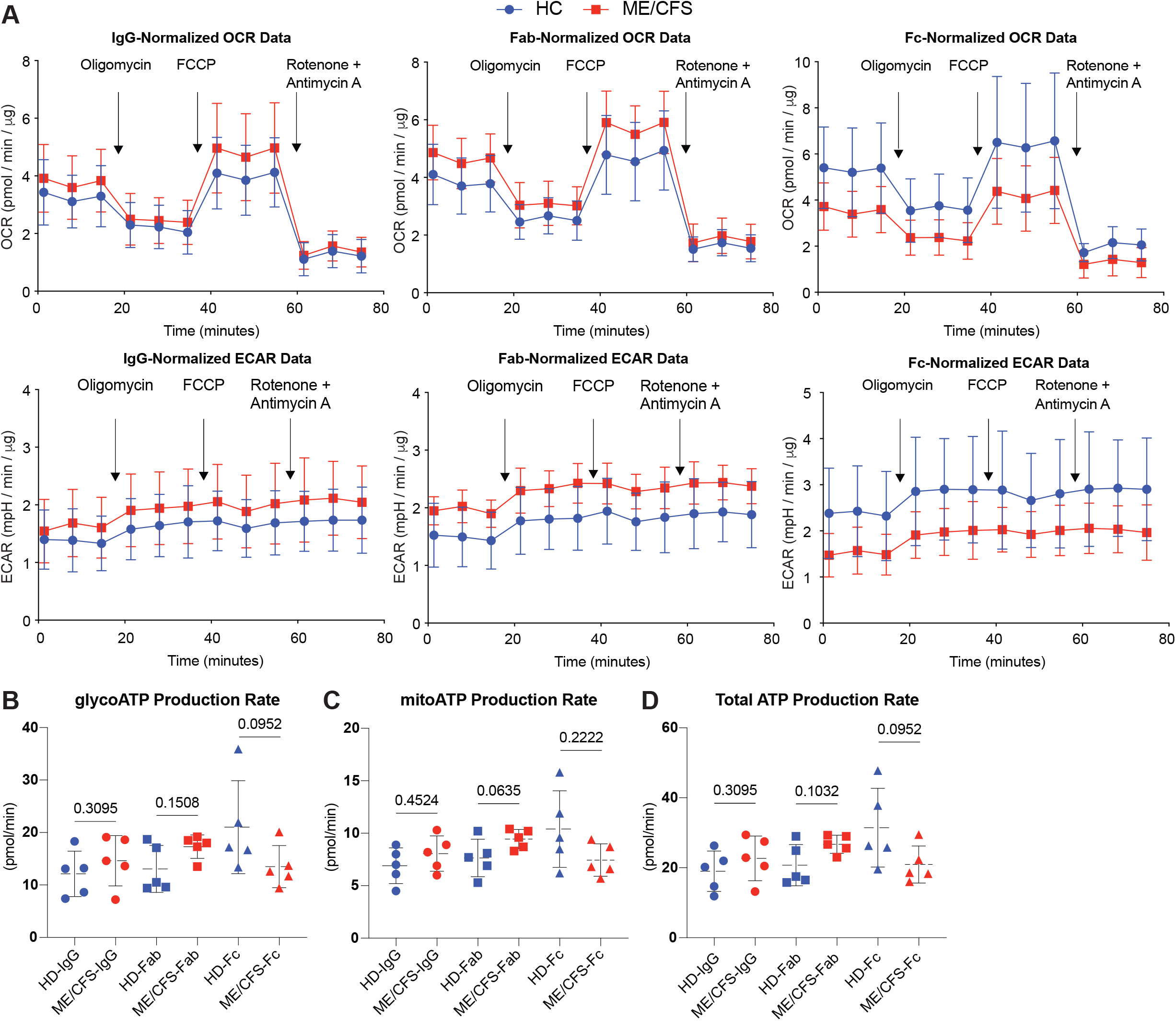
Fab and Fc fragments of IgG have variable effects on metabolic energetics in HUVECs. A. Normalized OCR and ECAR in HUVECs exposed to 1⍰μg/mL IgGs, Fab fragments, and Fc fragments from the same 5 ME/CFS and 5 healthy controls measured by Seahorse assay mito stress test. B-D. Seahorse real-time ATP rate assay in HUVECs exposed to 1⍰μg/mL IgGs, Fab fragments, and Fc fragments from the same 5 ME/CFS and 5 controls. B. Glycolytic ATP production rate. HD-IgG vs ME/CFS-IgG, ns P=0.3095. HD-Fab vs ME/CFS-Fab, ns P=0.1508. HD-Fc vs ME/CFS-Fc, ns P=0.0952. C. Mitochondrial ATP production rate. HD-IgG vs ME/CFS-IgG, ns P=0.4524. HD-Fab vs ME/CFS-Fab, ns P=0.0635. HD-Fc vs ME/CFS-Fc, ns P=0.2222. D. Total ATP production rate. HD-IgG vs ME/CFS-IgG, ns P=0.3095. HD-Fab vs ME/CFS-Fab, ns P=0.1032. HD-Fc vs ME/CFS-Fc, ns P=0.0952.

### IgGs from ME/CFS patients induce secretion of specific inflammatory cytokines

Alterations in mitochondrial structure and function can release various mitochondrial components and metabolic products that can function as damage-associated molecular patterns (DAMPs) and promote inflammation when released into the cytosol (*36*). Alternatively, direct interaction of IgG-TLR complexed ligands can induce pro-inflammatory cytokines (*37*). Therefore, we tested whether passive transfer of IgG to healthy PBMCs can trigger inflammatory markers often associated with the ME/CFS (Figure 5A). Ten well-known cytokines were measured in a multiplexed bead-based assay. The same IgGs were exposed to two separate PBMC samples from independent healthy donors, and two independent biological replicates were carried out for each IgG treatment. IgGs from a large subset of ME/CFS patients induced the secretion of four cytokines, including IL-1β (P=0.0460, IFN-γ (P=0.0675), TNFα (P=0.1653) and IL-6 (P=0.0675), (Figure 5B). However, this inflammatory cytokine induction phenotype was not uniform for all ME/CFS patients, making the differences statistically insignificant (Figure 5B) except for IL-1β. Notably, 48-hour exposure of PBMCs to IgG from MS patients significantly induced the levels of 3 cytokines, including IL-5, IL-10, and TNFβ, and reduced levels of TGFα (Figure 5B). A similar trend was also observed when PBMCs were treated with IgG from PCS-CFS patients. However, IgGs from ME/CFS patients had no effect on the secretion of these cytokines. Levels of IFNα2 was not altered by IgG from all 4 study groups. We verified alterations in some of these cytokines using alternative ELISA-based commercial assays (Supplementary Figure S2A) that mostly replicated the multiplex assay results. In these studies, IgGs from ME/CFS patients showed a contrasting phenotype to those from PCS-CFS patients. In summary, these results demonstrated that IgG from patients can induce the secretion of specific inflammatory cytokines; however, they could not establish a direct causal link between IgG-induced mitochondrial fragmentation and inflammation, suggesting a role for a mitochondria-independent pathway.

**Figure 5.**
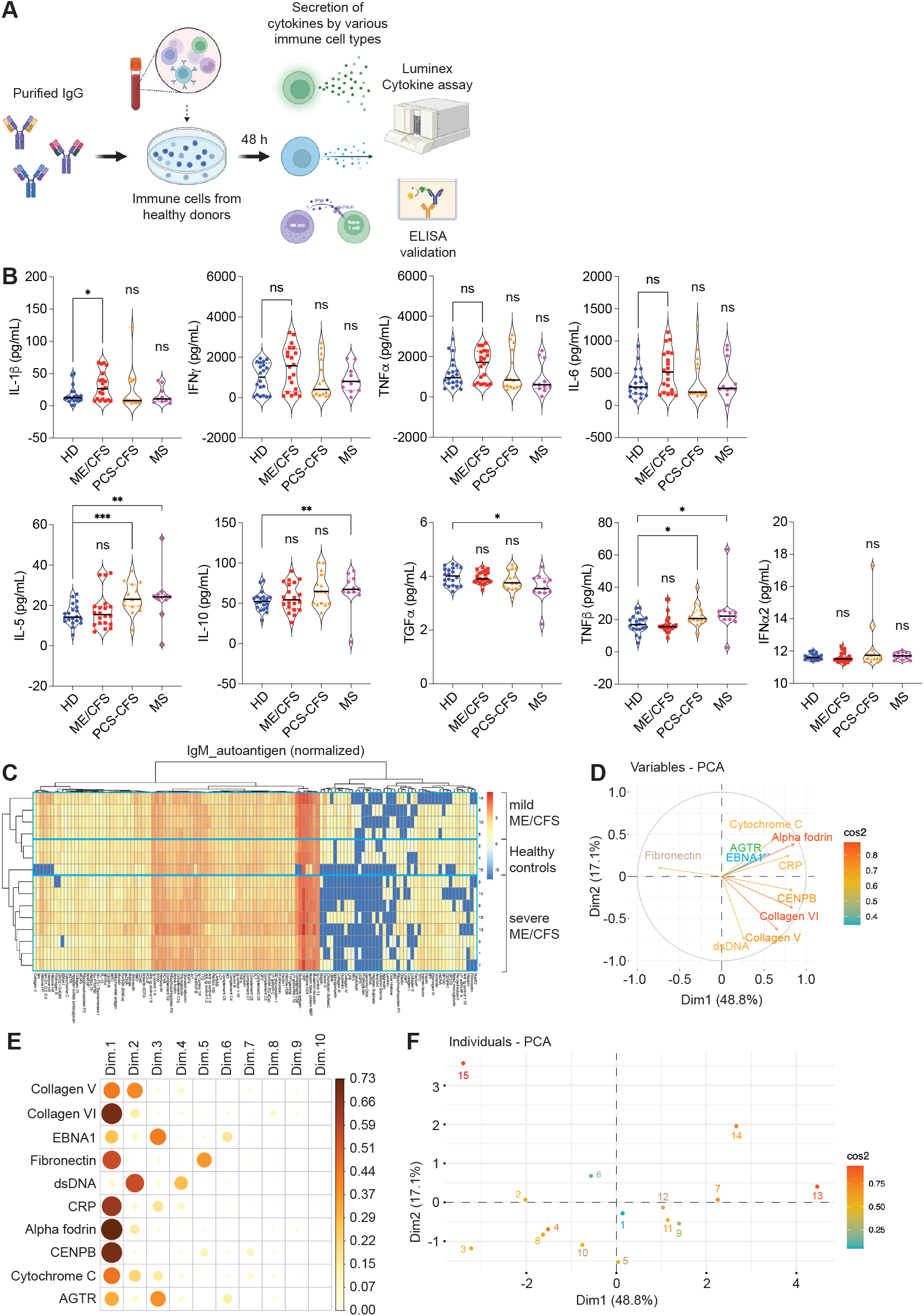
IgG induces inflammation and autoantibody signature in ME/CFS. A. Schematics of experimental setup to measure inflammatory cytokines produced by IgG-induced inflammation. B. Violin plots showing results of multiplex bead-based assays to measure secreted inflammatory cytokines from PBMCs of two healthy donors after 48 hours of exposure of PBMCs to IgG from ME/CFS (n=20), PCS-CFS (n=15), MS (n=11), and healthy donors (n=20). Two-tailed Mann-Whitney U test. (IL-1β) HD vs ME/CFS, * P=0.0460. HD vs PCS-CFS, ns P=0.4785. HD vs MS, ns P=0.7132. (INF-*γ*) HD vs ME/CFS, ns P=0.0675; HD vs PCS-CFS, ns P=0.9345; HD vs MS, ns P=0.8550. (TNF-α) HD vs ME/CFS, ns P=0.1653. HD vs PCS-CFS, ns P=8307. HD vs MS, ns P=0.1566. (IL-6) HD vs ME/CFS, ns P=0.0675; HD vs PCS-CFS, ns P=0.9345; HD vs MS, ns P=0.9514. (IL-5) HD vs ME/CFS, ns P=0.8201, *** P=0.0005; HD vs MS, ** P=0.0030. (IL-10) HD vs ME/CFS, ns P=0.4612; HD vs PCS-CFS, ns P=0.0545; HD vs MS, ** P=0.0016. (TGFα) HD vs ME/CFS, ns P=0.7329. HD vs PCS-CFS, ns P=0.2302. HD vs MS, * P=0.0144. (TNF-β) HD vs ME/CFS, ns P=0.6783; HD vs PCS-CFS, * P=0.0143; HD vs MS, * P=0.0202. (IFNα2) HD vs ME/CFS, ns P=0.1803. HD vs PCS-CFS, ns P=0.5366. HD vs MS, ns P=0.7838. C. Multivariate analysis of clusters based on distance metrics derived from IgM antibody levels for a panel of autoantigens. Log-transformed scaled data showing relative differences between different variables in both healthy controls and patients. D. The Variables Factor map for the Principal Components (combining data from patients and healthy controls) shows the projection of the top 10 Autoantigen variables onto the plane spanned by the first two Principal Components. E. The Contribution map shows the contributions of the top 10 Autoantigens to the top 10 dimensions. F. Biplot of PCA analysis showing the plot of individuals for the PCA in the Extended Data Figure 3b.

### Autoantibodies as a potential cause of mitochondrial fragmentation

IgG-induced mitochondrial fragmentation can be caused by specific autoantibodies enriched in ME/CFS and PCS-CFS patients. Numerous publications have shown evidence of one or more autoantibodies being increased in ME/CFS patients, making it challenging to correlate a specific autoantibody with the mitochondrial phenotype. We conducted a proof-of-concept pilot study utilizing commercial protein microarrays. To test potential IgG and IgM responses against selective autoantigens frequently involved in autoimmune diseases, we measured IgG and IgM levels against 120 autoantigens (Supplementary Table 2) in a small cohort of mild to severe ME/CFS patients (n = 12) and healthy controls (n = 3). Samples were blinded throughout the experimental procedure and data analysis. Multivariate clustering of log-transformed data revealed that patients could be separated into distinct groups, comprising healthy, mild/moderate, and severe patients (Figure 5C), based on IgM antibody levels against antigens such as PCNA, collagen V and VI, complement C3, and CRP (Figures 5D-5F). IgM against fibronectin was negatively associated with disease severity. A similar analysis of IgG levels against autoantigens was unable to differentiate the cases from the controls (Supplementary Figure S2B). One major limitation of our current approach, Sepharose G bead-based IgG purification, is that it does not purify the majority of natural IgM, which are pentameric structures, making it challenging to correlate IgM autoantibody levels to mitochondrial fragmentation. While these results suggest that IgM levels against known autoantigens can distinguish between ME/CFS disease groups and healthy individuals, they may not be responsible for mitochondrial fragmentation.

### Proteomic analysis of immune complexes

The immunoglobulin-mediated alterations in mitochondrial energetics could be transmitted through antigen-antibody complexes. ME/CFS is closely associated with immune dysregulation, yet no definitive biomarkers have been identified. Plasma proteomic analysis has been shown to be a powerful tool to identify biomarkers in certain diseases (*38, 39*), including ME/CFS (*40, 41*). Purified IgG from human serum mainly contains immunoglobulin fractions with freely available Fc receptors along with associated immune complex proteins. Unlike plasma proteomics, which analyzes all circulating proteins, immune complex proteomics enriches for antibody-bound proteins, making it specific for identifying immune complex-related proteins and disease-specific immune functions, such as those associated with autoimmune diseases. To identify fine changes within the immune complex compositions, we carried out mass spectrometry analysis of purified immunoglobulin fractions from ME/CFS (n = 40), PCS-CFS (n = 16), healthy controls (n = 39), and multiple sclerosis (n = 11) (Figure 6A). Our analysis did not detect any EBV or HHV-6A protein signature within the immune complex proteome. Based on the identified peptide signatures, we conducted a 3D principal component analysis (PCA) that nicely separated healthy controls from all three disease groups. Although there was no clear separation between the three disease groups, ME/CFS patients managed to cluster slightly further away from the PCS-CFS and MS groups (Figure 6B), as indicated by specific proteins that distinguished individual comparison groups (Figure 6C). A fold change analysis was conducted to identify the most significant up- and down-regulated proteins within the immune complex between different groups (Figures 6D-6G). Immune complexes from ME/CFS patients were enriched for several immunoglobulin heavy chain variable region genes (IGHVs), including IGHV1-58, IGHV4-8, IGHV3-16, IGHV3-66, and IGHV1-69-2 (Figure 6D). Interestingly, striated muscle-enriched protein kinase (SPEG), important for muscle maintenance, a kinase involved in muscle development and calcium handling, and Carboxypeptidase N Subunit 1 (CPN1) proteins, a metalloprotease a plasma metalloenzyme that regulates inflammatory peptides, were also increased within the immune complex of ME/CFS patients. One key protein involved in blood hemostasis, immune modulation, and vascularization, called Von Willebrand Factor (VWF) was significantly decreased within the immune complex of ME/CFS patients (Figures 6C, 6D). IGHV1-58 showed increased presence within the Immune complexes of PCS-CFS patients (Figure 6E) along with the protein kallikrein B (KLKB1). Similarly, IGHV3-16 was enriched within the immune complex of MS patients (Figure 6F).

**Figure 6.**
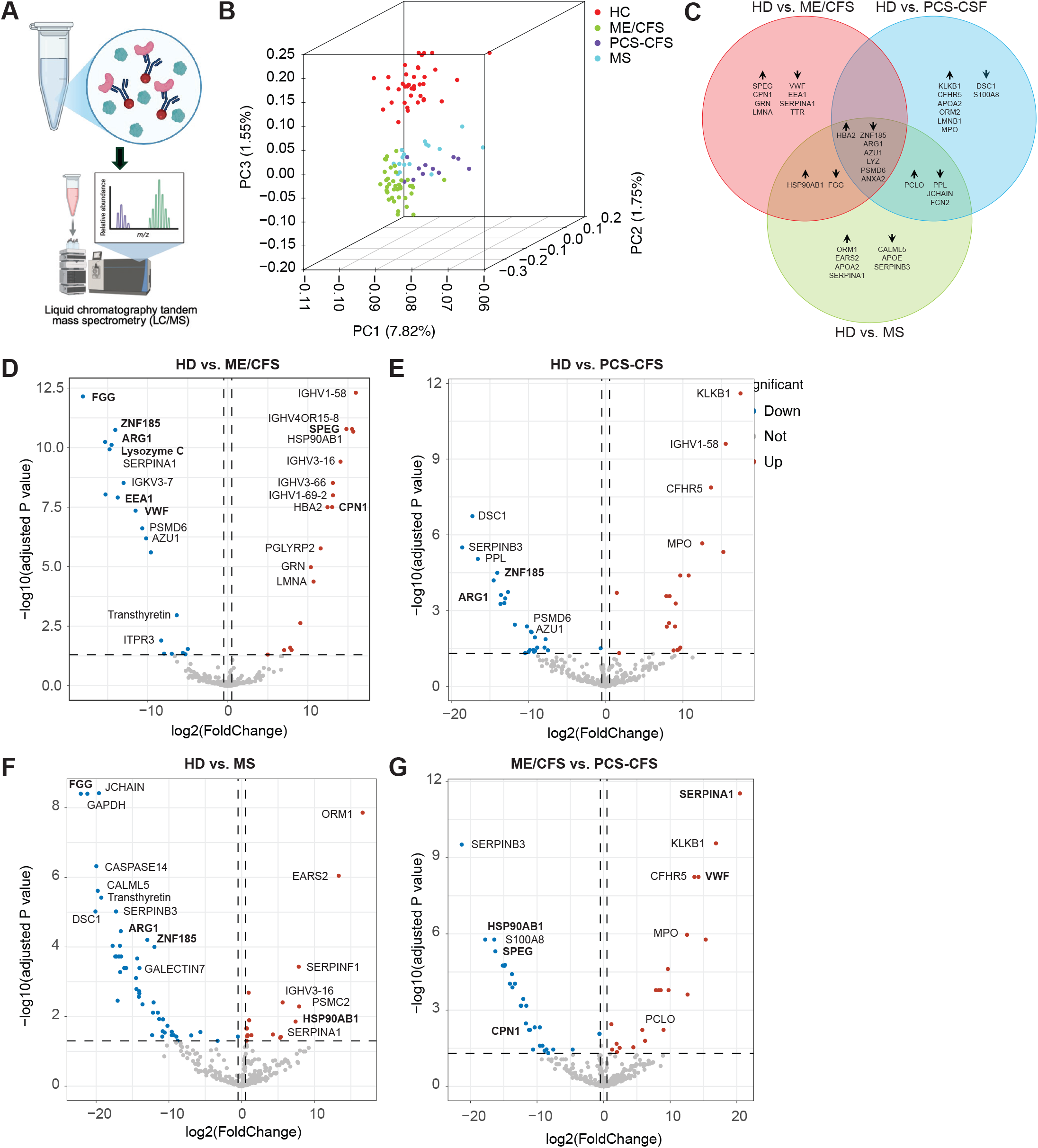
Immune proteome analysis shows distinct alterations in disease groups. A. Schematics of IgG-bound immune complex mass spectrometry-based proteomics. B. 3-Dimensional Principal Component Analysis (PCA) showing distribution of various disease groups and healthy controls on 3 different axis (PC1, PC2, and PC3). ME/CFS (n=39), PCS-CFS (n=15), MS (n=11) patients, and healthy controls (n=41). C. Venn diagram showing differentially detected proteins and commonly detected proteins within individual comparison groups as detected from mass spectrometry analysis of the immune proteome. D-G. Volcano plots of log_2_ fold changes and log_10_ adjusted P values for individual proteins detected by mass spectrometry. Top candidate genes upregulated (red) or downregulated (blue) with logFC > 1 and adjusted P value < 0.05 are indicated. Differentially detected proteins between healthy controls (HD) and ME/CFS (D), HD vs PCS-CFS (E), HD vs MS (F), and ME/CFS vs PCS-CFS (G).

We hypothesized that proteins that do not typically bind to immune complexes and are increased within the immune complexes of patients with a particular disease may represent proteins targeted by disease-specific autoantibodies (Figure 7A). Similarly, proteins that decrease within the immune complexes of a specific disease are potentially essential for the correct functioning and regulation of the immune system. These proteins may not be downregulated at the translational level. Instead, they may not be incorporated into the immune complex for some reason and, therefore, accumulate more in the serum and vice versa (Figure 7A). To validate this, we tested serum levels of several potential proteins that were either up- or down-regulated within the immune complex. KLKB-1 protein, which showed a significantly increased presence within the immune complex of PCS-CFS patients (Figure 6C), exhibited a trend of decreased presence in the total serum (P=0.126) (Figure 7B), but showed a significant decrease in the serum of ME/CFS patients (P=0.005). AZU1, which showed a reduced presence within the immune complex of all three disease groups (Figure 6C), exhibited a trend of increased presence in the serum of PCS-CSF patients (P = 0.115) (Figure 7B). VWF levels within the immune complex showed a trend of increased presence in the total serum of PCS-CSF patients (P=0.160) (Figure 7B). FCN2 and serotransferrin (TF) were detected in reduced amounts within the immune complex of all 3 disease groups and showed a significant increase in the serum of these patients (Figure 7C). The SPEG protein showed a comparative difference in its presence within the immune complex of HD and ME/CFS patients (Figures 6C, 6D); however, it was barely detectable in the serum of a few patients by ELISA, making validation difficult.

**Figure 7.**
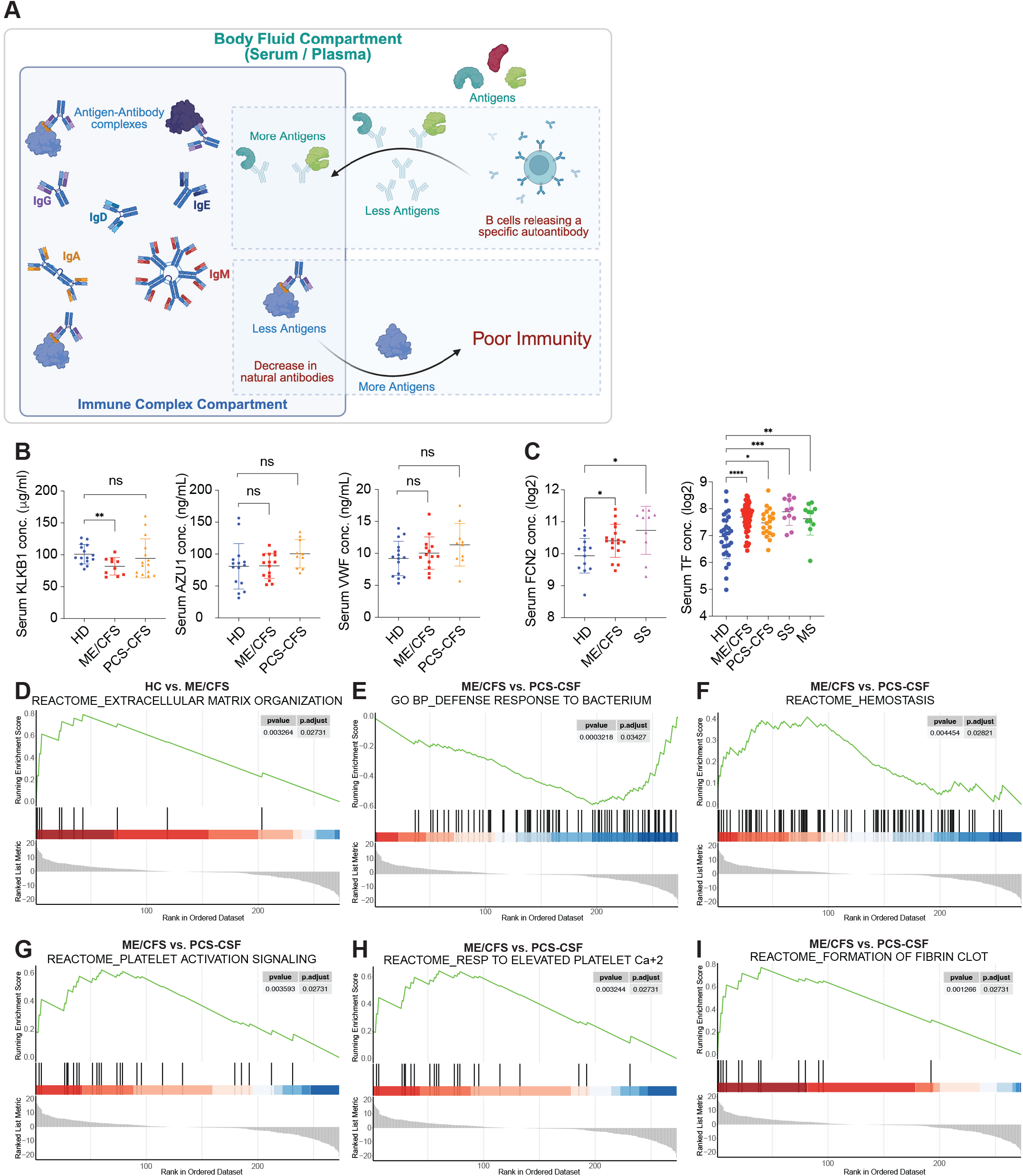
Gene Set Enrichment Analysis (GSEA) identified key altered pathways in patients with ME/CFS. A. Schematics showing potential alterations within antigen-antibody complexes and their effect on immunity. B. Serum KLKB1, AZU1, and VWF levels in healthy controls, ME/CFS, and PCS-CFS patients. Two-tailed Mann-Whitney U test. (KLKB1) HD vs ME/CFS, **P=0.0046. HD vs PCS-CFS, ns P=0.1286. (AZU1) HD vs ME/CFS, ns P=0.5123. HD vs PCS-CFS, ns P=0.1054. (VWF) HD vs ME/CFS, ns P=0.3705. HD vs PCS-CFS, ns P=0.1642. C. Serum FCN2 and TF levels in healthy controls, ME/CFS, PCS-CFS, Systemic Sclerosis (SS) and Multiple Sclerosis (MS) patients. Two-tailed Mann-Whitney U test. (FCN2) HD vs ME/CFS, *P=0.0287. HD vs SS, *P=0.0104. (TF) HD vs ME/CFS, ****P<0.0001. HD vs PCS-CFS, *P=0.0191. HD vs SS, ***P=0.0008. HD vs MS, **P=0.0087. D. GSEA Reactome analysis of the immune proteome identified enhanced extracellular matrix organization in ME/CFS patients compared to healthy controls. E. Gene Ontology (GO) analysis of biological processes in the immune proteome identified a decreased defense response to bacteria in PCS-CFS patients compared to ME/CFS patients. F-I. GSEA Reactome analysis of the immune proteome identified increased hemostasis (F), platelet activation and signalling (G), response to elevated platelet-derived cytosolic Ca^+2^ (H), formation of fibrin clot (I) in PCS-CFS patients compared to ME/CFS patients.

We performed gene set enrichment analysis (GSEA) using the immune complex proteome data. Only one significant positive enrichment for extracellular matrix reorganization was observed in the GSEA Reactome analysis of the immune proteome in ME/CFS patients compared to healthy controls (Figure 7D). No significant enrichment was observed in the immune proteome of PCS-CSF and MS patients. However, several significant differences were observed when the immune proteome of ME/CFS patients was compared to PCS-CSF. Gene Ontology (GO) analysis of biological processes showed a decreased response to bacterial infection in PCS-CSF patients (Figure 7E). GSEA Reactome analysis revealed increased hemostasis (Figure 7F), platelet activation (Figure 7G), platelet Ca2+ response (Figure 7H), and fibrin clot formation (Figure 7I) in the PCS-CSF patient immune proteome compared to ME/CFS. These results suggest specific alterations within the immune proteome of ME/CFS patients separate them from healthy controls and PCS-CFS patients.

## DISCUSSIONS

Mitochondrial phenotypes are cell- and tissue-specific. For example, among the nine cell types that circulate in the blood, 16 mitotypes with cell-specific mitochondrial proteomes and distinct mitochondrial phenotypes have been characterized (*42*). Non-circulating cells in the brain, heart, liver, kidneys, skeletal muscle, and other tissues also contain specialized mitochondria tailored to meet the metabolic and bioenergetic needs of the more than 400 different cell types in the body (*43*). Each cell type also has a characteristic copy number of mitochondrial DNA (mtDNAcn). Among circulating cells in the blood, B cells have the highest mtDNA copy number of 451 copies per single-copy nuclear gene, while neutrophils and NK cells have the lowest, with just 128. Among non-circulating cells in tissues, the range of mtDNAcn is broad, extending from over 6,700 copies per cell in highly metabolically active hippocampal neurons (*44*) down to just 115 copies per cell in skin fibroblasts (*45*). However, mitochondrial DNA encodes just 13 proteins of the 1,136 mitochondrial proteins that have been catalogued (*46*). The proteome of each mitochondrion is tailored to meet the specialized metabolic and bioenergetic needs of each differentiated cell type. Each of these parameters, i.e., mtDNA copy number, the expression of nuclear genes, and the tailored import of proteins into mitochondria, is adaptive. Under conditions of extreme stress, injury, or infection, the expression of up to 5,554 genes is regulated to contain, survive, and recover from the threat (*47*). The cell danger response (CDR) coordinates the regulation of all these genes and metabolic pathways in proportion to the degree of threat (*48*), and according to the timing after the threat has passed, needed to optimize healing and recovery (*49*). If healing is incomplete, the CDR is arrested and produces mitochondrial dysfunction and the symptoms of chronic illness. These symptoms define several multisystem chronic fatigue syndromes like ME/CFS and long COVID.

Hundreds of signals in the blood are used to coordinate the CDR in cells at remote distances from the original stress. These signals originate from specialized cells at the site of stress, injury, or infection, travel through the blood, and stimulate specialized functions in distant cells via cell-specific receptors. Earlier experiments from our group showed that signals released from a stressed cell growing in the lab were released into the medium and could be adoptively transferred to a healthy responder cell. In this way, signals from one cell type triggered the expression of previously silent HHV-6 miRNA (miR-aU14), mitochondrial fragmentation, a decrease in intracellular ATP (iATP), and a potent antiviral phenotype in the responding cells (*10*). In those experiments, the signaling cell type was not one that could make immunoglobulin. Therefore, we know that some of the signals capable of fragmenting mitochondria are IgG-independent. In patients with ME/CFS, hundreds of different cell-specific and non-specific cellular mediators are present in the blood. Some of these include metabolites such as extracellular ATP (eATP) and xanthine, which are universally produced in response to cellular stress and utilized in purinergic signalling (*50*). Markers of dysregulated mitochondrial fatty acid oxidation, like acylcarnitines, occur in ME/CFS (*19*). Signaling lipids like sphingomyelins, ceramides, eicosanoids, steroid hormones, bile acids, organic acids, and fatty acids are also dysregulated (*18*). Other mediators include cytokines such as TGF-β, (*51*) as diverse extracellular vesicles and exosomes (*52*). MicroRNAs are also dysregulated in ME/CFS and differ according to symptom severity (*53*). Even intact mitochondria are found in the blood at varying levels in healthy controls and patients with various acute and chronic medical conditions, and have been shown to be taken up and alter the metabolic phenotypes of the receiving cell types (*54*).

In this paper, we have focused on the role of a class of immunoglobulins purified by protein G affinity chromatography, IgG. We demonstrate that both the whole molecule and the Fab fragment of IgG act acutely to enhance the stress ATP response in treated cells. In contrast, the Fc fragment of IgG works chronically via Fc receptor (FcgR) internalization and recycling to inhibit the aerobic functions of mitochondria and induce a hypometabolic response in the responding cell types. Although classically considered to be restricted to immune cells like monocytes, macrophages, T cells, B cells, and NK cells, endothelial cells like the ones used in this study, also express FcgRs and are key players in the pathogenesis of ME/CFS (*55*). Several studies have suggested a potential role for autoantibodies in mitochondrial dysfunction (*56, 57*). IgG from patients with liver diseases significantly inhibited ADP-dependent mitochondrial respiration in oxidative muscle cells by targeting sarcomeres and thus disrupting the interaction between sarcomere and mitochondria (*56*). Here, we used purified IgGs from healthy controls, ME/CFS, PCS-CFS, and MS patients, and exposed them to HUVECs. We observed quantifiable changes in mitochondrial morphology (Figure 2A), with significantly more fragmented mitochondria in HUVECs exposed to IgGs from ME/CFS and PCS-CFS patients (Figures 2B, 2C). Moreover, IgG from female patients had a tendency to fragment mitochondria more effectively than IgG from male patients. This suggests that IgGs from ME/CFS and PCS-CFS patients can directly influence mitochondrial dynamics and correlate with the gender differences observed in these diseases. However, we did not observe a correlation between IgG-induced mitochondrial fragmentation and disease severity. Notably, this effect was not observed with IgGs from MS patients (Figures 2B, 2C), suggesting that mitochondrial fragmentation is not a general feature of autoimmune conditions but may involve disease-specific autoantibodies. Further analysis revealed that this mitochondrial fragmentation is not caused by excessive mitochondrial fission, which is typically mediated by Drp1. Instead, we observed a general reduction in multiple mitochondrial proteins, including Drp1, PLD6, Miga1, and mitofilin (Figure 2D), indicating potential mitochondrial destabilization and degradation.

In our study, IgGs from ME/CFS patients induced distinct metabolic energetics, which likely connect to the mitochondrial structural changes observed earlier. IgGs from ME/CFS patients led to increased mitochondrial respiration and glycolytic activity (Figure 3A), suggesting that the endothelial cells were under a high energy demand in the presence of IgG, a possible indication of cellular stress. To assess mitochondrial function independent of glycolysis, we replaced glucose with galactose in the assay medium, which forces cells to rely exclusively on OXPHOS. The higher spared respiration capacity induced by IgGs from ME/CFS was not displayed anymore in the presence of galactose (Figure 3E), suggesting glycolytic compensation as a primary source of stress in ME/CFS. Another study has already reported a significant increase in ATP production in PBMCs from ME/CFS, which primarily originates from non-mitochondrial respiration, such as glycolysis (*58*). Here, we demonstrate that IgGs from ME/CFS exhibit higher glycolysis compensation abilities. Elevated glycolysis could potentially lead to oxidative stress, increased ROS production (*16*), and impaired energy metabolism, ultimately contributing to mitochondrial dysfunction (*59, 60*). Interestingly, IgGs from PCS-CFS patients did not alter the mitochondrial energetics to the same level as those from ME/CFS patients. Overall, ME/CFS IgGs induced mitochondrial fragmentation and metabolic stress, without necessarily impairing oxidative phosphorylation (OXPHOS) function.

The targets of autoantibodies are often informative about the pathophysiology of autoimmune diseases. A distinguishing feature of autoantibodies is whether they target intracellular or extracellular proteins. Autoantibodies against extracellular targets can directly contribute to the disease progression by interfering with the functions of cell surface receptors or triggering the complement cascade, resulting in inflammation and tissue damage (*37, 61*). Our results support this role of IgGs, as the secretion of inflammatory cytokines, such as IL-5, IFN-γ, and TNF-β, was induced in healthy PBMCs through patient IgG. Although autoantibodies against intracellular proteins are generally considered unable to access their antigens in intact cells, disease pathogenesis may arise indirectly through immune-mediated mechanisms triggered by the antigen–antibody interaction (*62-64*). There has been limited studies focused on autoantibody penetration into cells. Here, we show a cell-type-specific IgG internalization in HUVECs. This internalization of IgG in HUVECs could potentially be caused by endocytosis, as indicated by the localization of internalized IgG within vesicle-like, double-membrane structures. However, it is crucial to understand the precise mechanism of this internalization. Our results showed decreased IgG internalization upon Fc receptor blockade, suggesting that the endocytosis is mediated by the neonatal Fc receptor (FcRn). FcRn is a potential candidate and known to facilitate IgG homeostasis and transport, and its expression in endothelial cells has been well studied. FcRn is localized both at the cell surface and within intracellular vesicles. Furthermore, the expression of FcRn is upregulated after IgG exposure in HUVECs (*65*). FcRn binds to the Fc region of IgGs in a pH-dependent manner, which subsequently protects them from lysosome degradation, thereby extending their half-life (*66*). This also allows the transport of IgGs across tissues such as the endothelium and epithelium (*67-69*). Besides the FcRn-mediated internalization on the cell surface, IgGs can also be taken up by a non-specific mechanism in endothelial cells, such as macropinocytosis or fluid-phase endocytosis. Within the endosome, IgG binds to FcRn, allowing it to be salvaged from lysosome degradation and recycled back to the cell surface (*70*). This process could become pathogenic when large amounts of autoantibodies are present in the circulation system. Taken together, these results demonstrate that IgG entry into cells is cell type-specific and likely mediated by distinct protein interactions involving both the Fab and Fc regions. Moreover, this entry can be partially inhibited by blocking the Fc receptor.

It is difficult to completely block the function of Fc receptors during live cell culture. Hence, we cannot conclude that Fc receptor-mediated IgG internalization is solely responsible for the IgG internalization observed in our results. After digesting the purified IgGs into Fab and Fc fragments, we observed that not only could the Fc fragments enter HUVECs, but the Fab fragments could also enter HUVECs. This suggests that multiple mechanisms may contribute to IgG internalization in HUVECs. Unlike Fc fragments, which could be potentially internalized via receptor-mediated mechanisms, Fab fragments lack natural receptors. However, it has been reported that Fab fragments can bind to cell surface antigens and thus be internalized, such as tumor antigens HER2 and EGFR (*71, 72*). Alternatively, in our case, Fab fragments are possibly internalized through non-specific, and endocytosis. However, without the Fc region, the Fab fragments are unable to interact with Fc receptors and may remain trapped within endosomes or in the cytoplasm. Further evidence supporting this hypothesis comes from the observed levels of internalization of IgGs, Fab fragments, and Fc fragments. Despite exposing HUVECs to equal amounts of IgGs, Fab fragments, and Fc fractions, the internalization levels varied without a clear pattern among the samples. However, the internalization of total IgGs and Fc fragments showed sample-dependent variation, while Fab fragment internalization was more evenly distributed across samples. This supports the hypothesis that the Fab fragments internalization occurs via a non-specific mechanism, while Fc receptor-mediated internalization is receptor-specific.

These findings highlight a complex and multifactorial mechanism of IgG internalization into endothelial cells. This is particularly important in autoimmune diseases and in the context of ME/CFS. The release of autoantibodies into endothelial cells could contribute to the endothelial and vascular dysfunction observed in ME/CFS. Although Fc receptors play an essential role in the uptake of IgGs, our data strongly suggest that another unspecific and non-receptor-mediated pathway contributes to this phenotype. The fact that both digested Fab and Fc fragments can enter HUVECs independently suggests that other classes of antibodies, such as IgM, IgA, or IgE, may also contribute to disease progression caused by autoantibodies. Moreover, the differential degradation patterns of IgGs observed between healthy controls and ME/CFS indicate that autoantibodies may persist longer intracellularly. Taken together, our results suggest that already well-studied mechanisms of cellular handling of IgGs may be potential factors in the pathophysiology of ME/CFS and other autoimmune diseases.

Immune complex proteomics specifically isolates antibody-bound proteins, enriching for antigens, autoantigens, and immune mediators. While plasma proteomics provides a broader overview of circulating proteins, immune complex proteomics offers a more targeted and enriched assay for studying autoimmune diseases, where immune activity plays a dominant role in disease progression and treatment outcomes (*73*). Our immune proteome analysis revealed specific changes within the immune complex of ME/CFS patients compared to healthy controls and other disease controls, which may be relevant to many of the clinical features of ME/CFS.

We observed increased SPEG protein amounts within the immune complex of ME/CFS patients (Figures 6C, 6D), suggesting the presence of autoantibodies against SPEG in ME/CFS. SPEG are muscle-expressed protein kinases. They play an essential role in muscle differentiation and maintenance (*74*). Loss of SPEG results in structural muscle defects, including mislocalization of focal adhesion proteins and abnormal calcium handling, which could potentially contribute to the pathophysiology in ME/CFS (*75*). SPEG is mostly intracellular, found in cardiac and skeletal muscle cells (*74*). It is unlikely to be found in significant amounts in serum samples. We were unable to detect sufficient amounts of SPEG in serum samples using ELISA. However, in the event of muscle or cardiac injury, SPEG might be released into serum (*76*), which requires a more sensitive technique to detect and quantify. We demonstrated decreased presence of VWF proteins within the immune complexes of ME/CFS and concurrently validated their increase in the serum. VWF is primarily involved in hemostasis by mediating platelet adhesion and stabilizing coagulation Factor VIII to initiate clot formation (*77*). Besides, VWF binds to C1q and reduces phagocytosis and pro-inflammatory cytokine secretion by macrophages (*78*). Studies have shown VWF’s role in the regulation of blood vessel formation and vascular dysregulation (*79*). These features are also observed in ME/CFS (*80*).

Increased presence of KLKB1 (Figures 6C, 6D) within the immune complex suggested the presence of autoantibodies against this protein. KLKB1 is a serine protease precursor. Upon activation by Factor XII, it converts into plasma kallikrein, which plays a crucial role in coagulation, fibrinolysis, inflammation, and blood pressure regulation (*81*). Serotransferrin (TF) was detected in increased amounts within the immune complexes of a small subset of ME/CFS and PCS-CFS patients, making it statistically insignificant within the entire cohort. However, our results showed a significant increase in total TF amounts in the serum (Figure 6I). TF is a glycoprotein that binds and delivers iron to various tissues and maintains the iron homeostasis (*82*). Although no study has yet described the role of TF in immune complexes, TF has been shown to play a crucial role as an immune regulator. During an ongoing bacterial infection, TF inhibits bacterial growth by restricting iron from pathogens and thus reduces the inflammation (*83*). Our results show that alterations in TF amounts in serum may not be a characteristic feature of ME/CFS. Rather, it seems to be an overlapping feature of many autoimmune diseases.

We detected several key proteins, including AZU1, ARG1, ZNF185, and LYZ, in reduced amounts within the immune complexes of all three patient types (Figure 6C), suggesting their potential role in immune dysfunction and autoimmunity. We validated an increase in serum AZU1 in both ME/CFS and PCS-CFS (Figure 6H). AZU1, an antimicrobial protein secreted by neutrophils, can be incorporated into small extracellular vesicles (SEVs), resulting in elevated calcium levels in endothelial cells and increased vascular permeability (*84*). It plays a crucial role in immune complex formation by activating monocytes and macrophages to enhance their ability to recognize and process immune complexes (*85*). Recent studies have suggested the negative impacts of AZU1 on mitochondrial function by reducing oxygen metabolism, decreasing mitochondrial membrane potential, and contributing to cellular stress and apoptosis (*86, 87*). ZNF185 prevents stress fiber formation through the inhibition of RhoA in endothelial cells(*88*). ZNF185 knockdown disrupts actin filaments and promotes stress fiber formation without inflammatory mediators. Constitutive activation of RhoA is induced by ZNF185 knockdown, which results in forskolin-resistant endothelial barrier dysfunction. Knockout of mouse Zfp185, which is an orthologous gene of human ZNF185 increases vascular leakage in response to inflammatory stimuli *in vivo*. ZNF185 is a p53 target gene following DNA damage (*89*). Upon genotoxic stress, caused by the DNA-damaging drug etoposide and UVB irradiation, ZNF185 expression is upregulated. In etoposide-treated cells, ZNF185 depletion does not affect cell proliferation and apoptosis but interferes with actin cytoskeleton remodeling and cell polarization. This makes ZNF185 a potential gene to focus in many autoimmune diseases.

We observed a decrease in SERPINA1 within the immune complexes of ME/CFS patients. However, we were unable to detect SERPINA1 in the serum by ELISA because of its low abundance. SERPINA1 is a serine protease inhibitor, particularly to neutrophil elastase, which inhibits excessive immune activation by suppressing neutrophil-driven inflammation, thereby protecting tissues from autoimmune damage (*90, 91*). However, increasing evidence has suggested SERPINA1 to be connected to mitochondrial dysfunction. Z mutation of SERPINA1 (ZAAT) leads to misfolded protein accumulation in the endoplasmic reticulum (ER) of hepatocytes (*92*). This misfolded ZAAT translocates to mitochondria for degradation and leads to abnormal mitochondrial morphology and function, dysregulated lipid metabolism, and increased ATP and reactive oxygen species (ROS) production (*93*).

The immune proteome analysis of our study groups revealed several significant alterations in pathways and biological processes. Only one significant alteration (extracellular matrix (ECM) organization) was predicted in ME/CFS patients through GSEA Reactome analysis (Figure 7A). In a previous study, we demonstrated a significantly increased level of fibronectin (FN1) in the serum of both ME/CFS and post-COVID patients, but not in MS patients (*94*). In this study, FN1 was detected in lower amounts within the immune complexes of a subset of patients. Alterations in ECM can lead to neuroinflammation(*95*), vascular dysfunction (*96*), mitochondrial dysfunction(*97*), fibrosis, and tissue stiffness (*98, 99*). The ECM is essential in maintaining vascular integrity and permeability. Dysregulation of the ECM could lead to capillary leakage, inadequate tissue perfusion, and altered mechanotransduction signaling in endothelial cells, resulting in orthostatic intolerance, postural orthostatic tachycardia syndrome (POTS), and other signs of microvascular dysregulation commonly observed in ME/CFS patients. The ECM may act as a regulatory interface among the immune system, nervous system, and vascular system in ME/CFS. Aberrant ECM remodeling could be a downstream effect of chronic inflammation, infection, or autoimmunity, and in turn, perpetuate the disease cycle. We did not observe similar pathway enrichments for PCS-CFS patients. However, several key differences were predicted between ME/CFS and PCS-CFS groups, including lower antibacterial defense, increased hemostasis, platelet activation and signaling, and fibrin clot formation (Figure 7B-7F). These pathways were enriched in the PCS-CFS groups compared to the ME/CFS group. All these processes are altered in post-COVID patients and have been an intense topic of study recently (*100*). Our results demonstrate the potential for developing disease markers based on the proteome of immune complexes. However, more data and in-depth studies are required to validate some of these markers.

SARS-CoV-2 infection-induced ME/CFS (PCS-CFS) patients share clinical features with ME/CFS patients with unknown etiology. Our study offers a biological perspective for comparing various molecular aspects of these two conditions. While IgG-induced mitochondrial fragmentation and alteration in energetics were more prominent in post-infectious ME/CFS patients, IgG-induced alterations in inflammation-associated cytokines were prominent in PCS-CFS patients. IgG proteomics also showed distinct signatures within the immune complex of these two groups. We hypothesize that PCS-CFS patients represent an early stage of ME/CFS, where inflammation dominates clinical outcomes, whereas ME/CFS patients represent the chronic nature of the disease, characterized by hypometabolism and consequent metabolic alterations that influence clinical presentation. In summary, our data provide a mechanistic basis for considering therapeutic strategies targeting autoantibody-mediated pathology.

## MATERIALS AND METHODS

### KEY RESOURCES TABLE

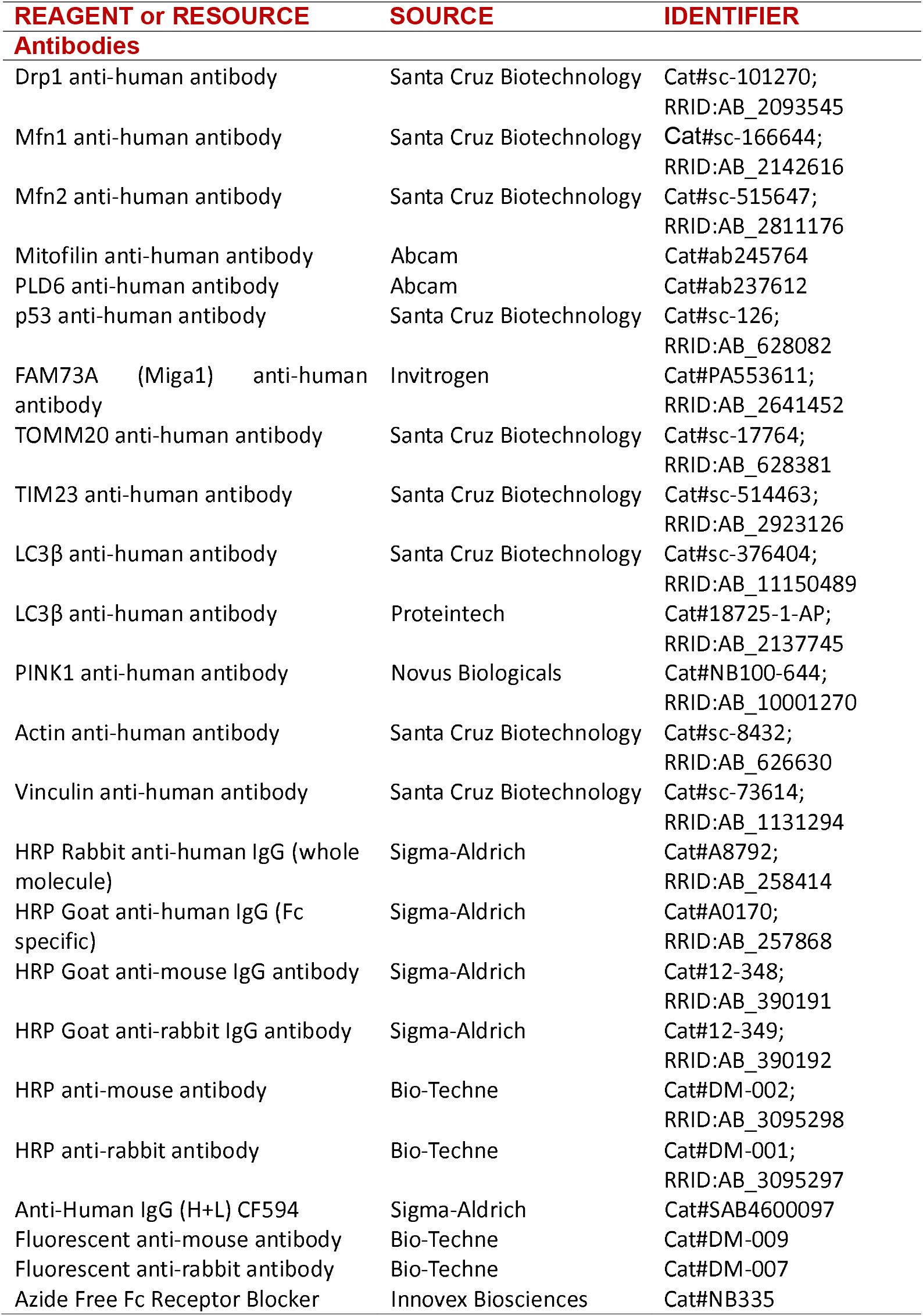
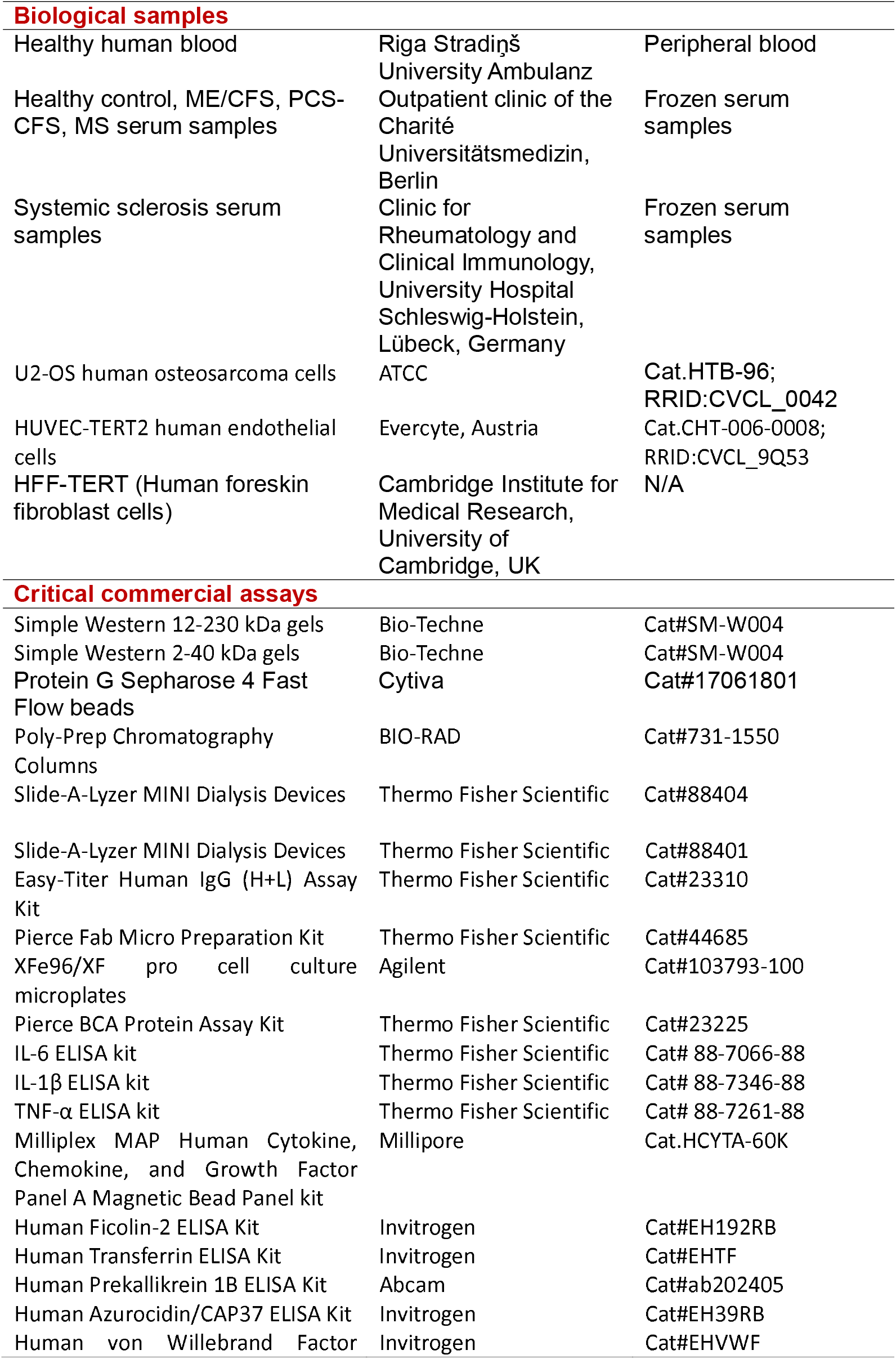
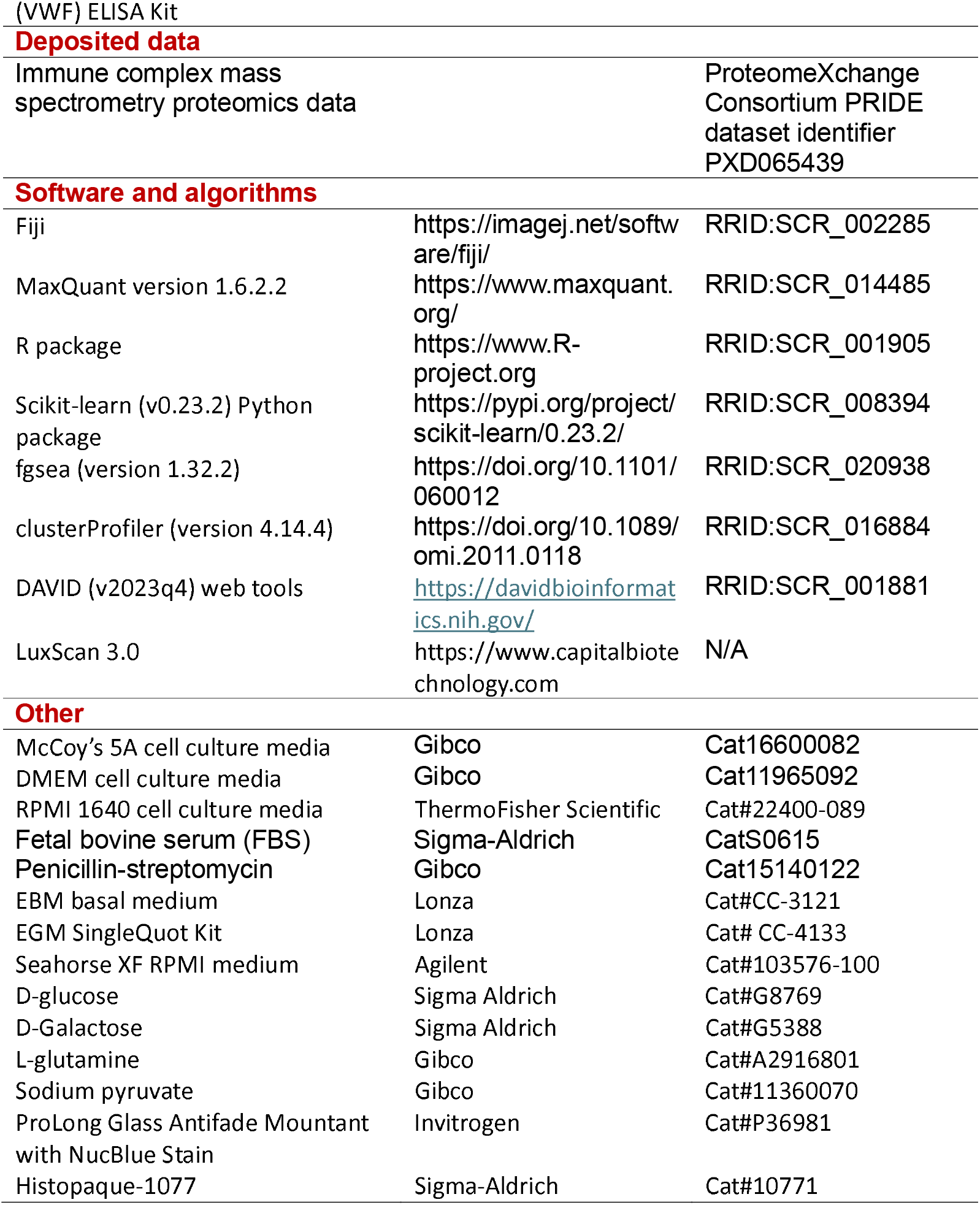

### RESOURCES AVAILABILITY

#### Lead contact

Further information and requests for resources and reagents should be directed to Bhupesh K Prusty.

#### Materials availability

This study did not generate new, unique reagents.

#### Data and code availability

The mass spectrometry proteomics data for the serum immunoglobulin proteome study have been deposited in the ProteomeXchange Consortium via the PRIDE partner repository with the dataset identifier PXD065439.

Any additional information required to reanalyze the data reported in this paper is available from the lead contact upon request.

This paper does not report any original code.

### EXPERIMENTAL METHODS AND SUBJECT DETAILS

#### Patient recruitment and serum collection

Postinfectious ME/CFS, including post-COVID ME/CFS (PCS-CFS) patients, relapsing-remitting multiple sclerosis (RRMS) patients, and gender- and age-matched healthy controls, were recruited at the outpatient clinic of the Charité Universitätsmedizin, Berlin, between 2020 and 2023. Diagnosis of ME/CFS was based on the 2003 Canadian Consensus Criteria and exclusion of other medical or neurological diseases that may cause fatigue by a comprehensive clinical and laboratory evaluation. RRMS patients have been suffering from fatigue relevant to everyday life and have to be free from relapses as well as from steroid treatment for at least 6 months. Biosamples of ME/CFS patients and healthy controls were collected by the group of Prof. Scheibenbogen, and biosamples of MS patients were collected by the group of Prof. Paul. The study was approved by the Ethics Committee of Charité Universitätsmedizin Berlin (EA2/067/20; EA2_066_22; EA4_174_22) in accordance with the 1964 Declaration of Helsinki and its subsequent amendments, as well as the approval from the Medizinische Ethikkommission of JMU, Würzburg (83/23). A total of 21 serum samples from patients diagnosed with systemic sclerosis were collected by the group of Prof. Riemekasten at the Clinic for Rheumatology and Clinical Immunology, University Hospital Schleswig-Holstein, Lübeck, Germany, as part of the same approved study. These samples were only used for potential biomarker validation. All donors gave informed consent. Whole blood samples from each subject were allowed to clot at room temperature and then centrifuged at 2000 x g for 10 min. The serum was stored in aliquots at -80°C. Patient demographic is presented in Supplementary Table 1.

#### Cell culture

U2-OS (HTB-96) cells were purchased from ATCC, and HUVEC-TERT2 cells (CHT-006-0008) were purchased from Evercyte, Austria. Human foreskin fibroblast cells (HFF-TERT) were kindly provided by Prof. Michael Weekes (Cambridge Institute for Medical Research, University of Cambridge, UK). U2-OS and HFF-TERT cells were grown in McCoy’s 5A (Gibco, 16600082) and DMEM media (Gibco, 11965092), respectively, supplemented with heat-inactivated 10% fetal bovine serum (FBS) (Sigma-Aldrich, S0615) and 1% penicillin-streptomycin (Gibco, 15140122). HUVEC-TERT2 cells were grown in EBM basal medium (Lonza, Cat#CC-3121) supplemented with Components of EGM SingleQuot Kit (Lonza, Cat# CC-4133: BBE, hEGF, hydrocortisone, ascorbic acid), 10% FBS, and 20 µg/ml G418. All cell lines were cultured at 37 °C with 5% CO2. Cells carrying stable GFP expression within mitochondria were developed as mentioned before (*101*). Cells stably expressing GFP were created by cloning the mitochondrial-targeted GFP into pLVTHM vector backbone and transducing target cells with the lentivirus as mentioned before (*101*). All the cell lines were frequently tested for Mycoplasma contamination and were authenticated by sequencing, wherever necessary.

#### Average mitochondrial surface area and mitochondrial number analysis

Software and a modified algorithm for mitochondrial size and number measurement were previously described by us in detail (*101, 102*). All image-processing and analysis steps were performed using Fiji (*103*).

#### Immunofluorescence Microscopy

Cells were seeded in 12-well plates on glass coverslips overnight and exposed to IgG. Cells were washed in PBS and fixed with 4% paraformaldehyde for 30 min. After washing, the cells were permeabilized using 0.2% Triton-X100 in PBS for 10 min and blocked with 10% FCS in PBS for 1h. Cells were then incubated for 1 h with Anti-Human IgG (H+L) CF594 (#SAB4600097, Sigma-Aldrich) in 2% FCS in PBS and washed three times in PBS. DAPI was used for staining cell nuclei. When GFP mitochondria and DAPI are the only channels required, the DAPI staining was followed directly after fixation. Samples were mounted onto slides using ProLong Glass Antifade Mountant with NucBlue Stain (#P36981, Invitrogen). Samples were analyzed on a confocal microscope (LSM 510 Meta, Carl Zeiss AG).

#### Immunoblotting

Immunoblotting was carried out as described before (*104, 105*) using antibodies: Drp1 antibody (#sc-101270, Santa Cruz Biotechnology), Mfn1 antibody (#sc-166644, Santa Cruz Biotechnology), Mfn2 antibody (#sc-515647, Santa Cruz Biotechnology), PLD6 antibody (#ab237612, Abcam), FAM73A Antibody (#PA553611, Invitrogen), p53 antibody (#sc-126, Santa Cruz Biotechnology), TOMM20 antibody (#sc-17764, Santa Cruz Biotechnology), TIMM23 antibody (#sc-514463, Santa Cruz Biotechnology), LC3β antibody (#sc-376404, Santa Cruz Biotechnology). Equal protein loading was confirmed by the following antibodies: Actin antibody (#sc-8432, Santa Cruz Biotechnology) and Vinculin antibody (#sc-73614, Santa Cruz Biotechnology). All primary antibodies were used at a dilution of 1:1,000 to 1:3,000. HRP-conjugated secondary antibodies, which are HRP Rabbit Anti-Human IgG (whole molecule) (#A8792, Sigma-Aldrich), Goat Anti-Human IgG (Fc specific) HRP (#A0170, Sigma-Aldrich), Goat Anti-Mouse IgG Antibody, HRP-conjugate (#12-348, Sigma-Aldrich) and Goat Anti-Rabbit IgG Antibody, HRP-conjugate (#12-349, Sigma-Aldrich), were used at a dilution of 1:2,000 to 1:10,000.

1 *μ*L of HUVEC protein lysates after IgG exposure were run in parallel on an automated capillary Simple Western (JESS from Bio-Techne) for proper quantification. Simple Western was performed according to the manufacturer’s protocol using 12-230 kDa (#SM-W004, Bio-Techne) or 2-40 kDa (#SM-W004, Bio-Techne) separation modules with chemiluminescent Anti-Mouse (#DM-002, Bio-Techne), Anti-Rabbit (#DM-001, Bio-Techne) or fluorescent Anti-Mouse (#DM-009, Bio-Techne), Anti-Rabbit (#DM-007, Bio-Techne) detection modules, and results were analyzed using Compass software (version 6.3.0). Assays were performed using antibodies: Mitofilin antibody (#ab245764, Abcam), Drp1 antibody (#sc-101270, Santa Cruz Biotechnology), TIMM23 (#sc-514463, Santa Cruz Biotechnology), PLD6 antibody (#ab237612, Abcam), FAM73A (Miga1) Antibody (#PA553611, Invitrogen), LC3β antibody (#18725-1-AP, Proteintech), PINK1 antibody (#NB100-644, Novus Biologicals). Equal protein loading was confirmed by Vinculin antibody (#sc-73614, Santa Cruz Biotechnology). All primary antibodies were used at a dilution of 1:50 to 1:500.

#### Immunoglobulin purification

150 µl Protein G Sepharose 4 Fast Flow (#17061801, Cytiva) was loaded onto Poly-Prep Chromatography Columns (#731-1550, BIO-RAD). After washing the beads with PBS, 500 µL of serum sample was loaded and passed through the column three times, which was followed by three washes with PBS. Protein G-bound serum IgG was eluted by Glycine pH 2.7 and then neutralised with 1M Tris-HCl pH 8.0. IgG elute was dialyzed against PBS in Slide-A-Lyzer MINI Dialysis Devices (#88404, Thermo Fisher Scientific). IgG concentration was measured by Easy-Titer Human IgG (H+L) Assay Kit (#23310, Thermo Fisher Scientific).

#### Fab fragments purification

Fab fragments of immunoglobulin G were purified using the Pierce Fab Micro Preparation Kit (#44685, Thermo Fisher Scientific), according to the manufacturer’s protocol. Fab fragments and flow-through containing digested Fc fragments were dialyzed against PBS in Slide-A-Lyzer MINI Dialysis Devices (#88401, Thermo Fisher Scientific). The concentrations of these fragments were quantified by NanoDrop™ 2000 Spectrophotometers (Thermo Fisher Scientific).

#### IgG exposure in cell culture

U2-OS or HUVEC-TERT2 cells carrying soluble mitoGFP were seeded on 6-well plates and cultured overnight. Cell culture medium was replaced by fresh medium containing 1 µg/ml serum IgG or 0.5 µg/ml Fab fragments or flow-through, allowing cells to be exposed to serum-derived IgG, Fab fragments, or flow-through containing Fc fragments. Cells were collected after 16 hours and 36 hours for Western blot or immunofluorescence experiments.

#### Fc receptor blocking assay

One hour prior to IgG exposure, the cell culture medium was replaced with fresh medium containing (20 μL) Azide Free Fc Receptor Blocker (#NB335, Innovex Biosciences). Following incubation, an equal volume of culture medium containing 2⍰µg/mL serum IgG was added to each well.

#### Seahorse Assay extracellular flux analysis

Mitochondrial function was assessed using the Seahorse XF Cell Mito Stress Test on the XFe96 Extracellular Flux Analyzer (Seahorse Bioscience). XFe96/XF pro cell culture microplates (#103793-100, Agilent) were pre-coated with 0.1% gelatin (#0646.1, Carl Roth) and seeded with 4,000 HUVECs per well. After overnight incubation, cells were exposed to purified serum IgG for 16 hours. All experimental conditions were performed in triplicate. On the day of the assay, the culture medium was replaced with Seahorse XF RPMI medium (#103576-100, Agilent), supplemented with 10⍰mM D-glucose (#G8769, Sigma Aldrich), 2⍰mM L-glutamine (#A2916801, Gibco), and 1⍰mM sodium pyruvate (#11360070, Gibco), and adjusted to pH 7.4. When required, 10 mM D-glucose was replaced to 10 mL D-Galactose (#G5388, Sigma Aldrich). Plates were incubated for 1 hour at 37⍰°C in a non-CO_2_ incubator prior to measurement. The Mito Stress Test was conducted according to the manufacturer’s instructions. Briefly, baseline oxygen consumption rate (OCR) and extracellular acidification rate (ECAR) was measured, followed by sequential injections of 1.5⍰*μ*M oligomycin (ATP synthase inhibitor), 1.0 μM FCCP (mitochondrial uncoupler), and 0.5⍰*μ*M rotenone combined with 0.5⍰*μ*M antimycin A (Complex I and III inhibitors, respectively). Each injection step included three measurement cycles consisting of mixing and measuring phases. Following the Seahorse assay, cells were lysed in 0.11M NaOH containing protease inhibitors and incubated at room temperature for 10 minutes. Protein concentration was quantified using Pierce BCA Protein Assay Kit (#23225, Thermo Fisher Scientific) according to the manufacturer’s protocol, for normalization of seahorse assay data. Seahorse assay data were recorded, processed, and analyzed using software Wave (Agilent). Mito Stress Test reports were generated according to the Seahorse XF Cell Mito Stress Test Report Generator template. OCR and ECAR values were analyzed to calculate key mitochondrial parameters, including basal respiration, ATP-linked respiration, proton leak, maximal respiration, spare respiratory capacity, and non-mitochondrial respiration. To quantify ATP production rates, the same Seahorse data set from the Mito Stress Test was reanalyzed using Seahorse XF Real-Time ATP Rate Assay Report Generator. OCR and ECAR values obtained before and after injection of oligomycin and rotenone/antimycin A were used to calculate mitochondrial ATP production, glycolytic ATP production, and total ATP production rates.

#### IgG-induced Inflammation Assay using human PBMCs

Peripheral blood was obtained from an apparently healthy adult donor under the ethical approval of Riga StradiņŠ University (RSU) Research Ethics Committee (2-PĒK-4/784/2025). PBMCs were isolated by density gradient centrifugation using Histopaque-1077 (Sigma-Aldrich, Cat# 10771) according to the manufacturer’s instructions. After centrifugation, the PBMC layer was collected, washed twice with sterile phosphate-buffered saline (Sigma-Aldrich, Cat# D8537), and resuspended in complete RPMI 1640 medium (ThermoFisher Scientific, Cat# 22400-089) supplemented with 10% heat-inactivated, filtered fetal bovine serum (FBS) and 1% Penicillin-Streptomycin.

Cells were seeded in flat-bottom 96-well plates (Sarstedt, Cat# 83.1835.300) at a density of 2 × 10^5^ cells per well in 200 µL of culture medium. Dialyzed IgG (final concentration: 50 µg/mL) from serum samples of healthy controls, patients with ME/CFS, PCS-CFS, or MS was added to the freshly isolated PBMCs immediately upon plating. Cells were incubated at 37⍰°C in a humidified atmosphere containing 5% CO_2_ for 48 hours. Following incubation, culture supernatants were collected and centrifuged at 400 × g for 5 minutes to remove cell debris. Concentrations of interleukin-6 (IL-6), interleukin-1β (IL-1β), and tumor necrosis factor alpha (TNF-α) were measured using commercially available enzyme-linked immunosorbent assay (ELISA) kits (ThermoFisher Scientific: IL-6, Cat# 88-7066-88; IL-1β, Cat# 88-7346-88; TNF-α, Cat# 88-7261-88) following the manufacturer’s protocols. Absorbance was read at 450 nm using a microplate reader (ThermoFisher, Varioskan LUX), and cytokine concentrations were calculated based on standard curves generated with recombinant standards provided in each kit.

#### Mass spectrometry sample preparation

A fraction of serum IgG-bound protein G beads from IgG purification was eluted by NuPAGE LDS Sample Buffer (4X) (#NP0007, Thermo Fisher Scientific). The eluates were sent for mass spectrometry.

#### Single-pot, solid-phase-enhanced sample preparation (SP3)

Samples were processed using an adapted SP3 protocol (*106*). Briefly, 200 µl reconstitution solution was added to each sample prepared in 50 µl NuPAGE LDS sample buffer (Life Technologies). Reduction was performed using 5 mM DTT, followed by alkylation with 20 mM iodoacetamide. 10 mM additional DTT was used for quenching. Equal volumes of two types of Sera-Mag Speed Beads (Cytiva, #45152101010250 and #65152105050250) were combined, washed with water, and 10 ⍰ µL of the bead mix were added to each sample. 260 µl 100% ethanol was added, and samples were incubated for 5 min at 24 °C, 1000 rpm. Beads were captured on a magnetic rack for 2 min, and the supernatant was removed. Beads were washed twice with 200 µl 80% ethanol (Chromasolv, Sigma) and then once with 1000 µl 80% ethanol. Digestion was performed on beads with 0.25 µg Trypsin (Gold, Mass Spectrometry Grade, Promega) and 0.25 µg Lys-C (Wako) in 100 µl 100 mM ammonium bicarbonate at 37 °C overnight. Peptides were desalted using C-18 Stage Tips(*107*). Each Stage Tip was prepared with three discs of C-18 Empore SPE Discs (3 M) in a 200 µl pipet tip. Peptides were eluted with 60 % acetonitrile in 0.1 % formic acid, dried in a vacuum concentrator (Eppendorf), and stored at -20 °C. Peptides were dissolved in 2 % acetonitrile / 0.1 % formic acid prior to nanoLC-MS/MS analysis.

#### NanoLC-MS/MS analysis

NanoLC-MS/MS analyses were performed on an Orbitrap Fusion (Thermo Scientific) equipped with a PicoView Ion Source (New Objective) and coupled to an EASY-nLC 1000 (Thermo Scientific). Peptides were loaded on a trapping column (2 cm x 150 µm ID, PepSep) and separated on a capillary column (30 cm x 150 µm ID, PepSep) both packed with 1.9 µm C18 ReproSil and separated with a 120-minute linear gradient from 3% to 30% acetonitrile and 0.1 % formic acid and a flow rate of 500 nl/min. Both MS and MS/MS scans were acquired in the Orbitrap analyzer with a resolution of 60,000 for MS scans and 30,000 for MS/MS scans. HCD fragmentation with 35 % normalized collision energy was applied. A Top Speed data-dependent MS/MS method with a fixed cycle time of 3 s was used. Dynamic exclusion was applied with a repeat count of 1 and an exclusion duration of 90 s; singly charged precursors were excluded from selection. The minimum signal threshold for precursor selection was set to 50,000. Predictive AGC was used with AGC a target value of 4×10^5^ for MS scans and 5×10^4^ for MS/MS scans. EASY-IC was used for internal calibration.

#### Enzyme-Linked Immunosorbent Assay (ELISA)

ELISA was performed using the following kits according to the manufacturers’ protocols: Human Ficolin-2 ELISA Kit (#EH192RB, Invitrogen), Transferrin ELISA Kit (#EHTF, Invitrogen), Human Prekallikrein 1B ELISA Kit (#ab202405, Abcam), Human Azurocidin/CAP37 ELISA Kit (EH39RB, Invitrogen) and Human von Willebrand Factor (VWF) ELISA Kit (#EHVWF, Invitrogen).

#### Antigen Microarray experimental setup

Microarray studies for IgG and IgM against autoantigens were carried out in collaboration with Creative Biolabs, USA. Frozen serum samples without a prior freeze-thaw cycle were used for the assay. Each serum was digested with DNase I for 30 min at room temperature on a shaker. For the control, no serum sample was added. Slides carrying antigens against 120 autoantigens (for details of the antigens, please see Supplementary Table 2) were blocked in 100 mL blocking buffer at room temperature for 30 min on a shaker. Afterwards, the slides were washed twice with PBST, each for 5 min. 90 ml PBST was added to each serum sample or control mix. Diluted samples were added to each well of the slide (100 μL each) and incubated at room temperature for 1 hour on a shaker. Slides were washed with PBST 100 ml/well for 5 min on the shaker. Subsequently, slides were washed with blocking buffer 100 ml/each well for 5 min on a shaker and then with PBST 100 ml/each well) for 5 min on a shaker. Anti-human IgM secondary antibody was diluted to 1:1000 in PBST, and 100 μL of secondary antibody was added to each well. Slides were incubated at room temperature for 1 hour on the shaker, washed three times with PBST 100 *μ*l/each well, 5 min on the shaker. Slides were then washed twice each, first with 45 ml PBS in a 50 ml tube for 5 min on the shaker and then with 45 ml nuclease-free water in a 50 ml tube for 5 min on the shaker. GenePix 4000B microarray systems were used to scan the slide. The 532 nm channel was used to scan Cy3 fluorescence, and the 635 nm channel was used to scan Alexa Fluor-647 fluorescence.

#### Multiplex bead-based assay for inflammatory marker measurements

The cell culture supernatant from PBMCs was centrifuged at 5000 g for 5 minutes to obtain a cell- and debris-free supernatant. TNFα, IFNγ, IL-5, IL-10, IL-6, IL-1β, IFNα2, sCD40L, TGFα, and TNFα levels in cell culture supernatants were quantified using the Milliplex MAP Human Cytokine, Chemokine, and Growth Factor Panel A Magnetic Bead Panel kit (HCYTA-60K, Millipore) according to the manufacturer’s protocol. Briefly, the samples were combined with marker-specific antibody-coupled magnetic beads in a 96-well plate and incubated overnight at 4 °C. Following incubation, the plate was washed on a magnet, followed by incubations with biotinylated detection antibodies and streptavidin-phycoerythrin. Finally, the plate was washed on a magnet, and beads were resuspended in xMAP Sheath Fluid Plus (40-50021, Luminex). The plate was read using the Luminex 200 instrument and xPONENT software, and the results were analyzed using the GraphPad Prism software by constructing a 7-point standard curve with a 5-parameter logistic curve-fitting method. Each plate included a background and two quality controls. Before reading the plate, the Luminex 200 instrument was calibrated and verified using the Luminex 100/200 Calibration and Performance Verifications kits (LX2R-CAL-K25 and LX2R-PVER-K25, Luminex). Each IgG was exposed to two separate biological replicates from each PBMC sample. Two such independent experiments were carried out using separate donor-derived PBMCs. The obtained results were combined, and the average values of each cytokine were used for the results.

### QUANTIFICATION AND STATISTICAL ANALYSIS

#### Mass spectrometry data analysis

Raw MS data files were analyzed with MaxQuant version 1.6.2.2 (*108*). A database search was performed using Andromeda, which is integrated into the utilized version of MaxQuant. The search was performed against the UniProt Human Reference Proteome database (Release 2024_3, UP000005640, 82518 entries), the UniProt EBV database (Release 2024_3, UP000272970, 55 entries), and the UniProt HHV-6A database (Release 2024_3, UP000009295, 99 entries). Additionally, a database containing common contaminants was used. The search was performed with trypsin cleavage specificity, allowing up to 3 miscleavages. Protein identification was under control of the false-discovery rate (FDR; <1% FDR on protein and peptide spectrum match (PSM) level). In addition to MaxQuant default settings, the search was performed against the following variable modifications: Protein N-terminal acetylation, Gln to pyro-Glu formation (N-terminal Gln), and oxidation (Met). Carbamidomethyl (Cys) was set as a fixed modification. LFQ intensities were used for protein quantitation (*109*).

The raw MaxQuant output files revealed expression data for 783 proteins. We focused on raw intensity values as well as label-free quantification (LFQ) intensity values for further analysis. The data processing and analysis work consisted of 4 components: Data processing, Clustering and PCA, Differential expression analysis, and pathway enrichment analysis. The first step of data processing involved contaminant filtering, removal of proteins with excessive missing values, imputation of missing values, and log transformation. We removed contaminants and low-quality proteins from the MaxQuant output data. Proteins filtered out included proteins with Potential contaminant value +, proteins with Reverse value +, and proteins identified only by site value +. Locally developed Python code was used for this purpose. To remove proteins with excessive missing values, we removed proteins with raw intensity values equaling 0 for at least 70% of the samples within the group. Missing values were evaluated using plots generated with the Amelia (v1.8.1) R package’s missmap() function, with default parameters. Imputation was performed on LFQ intensity data, although proteins with missing values were identified based on raw intensity. Specifically, when the raw intensity value for a protein was 0, it was considered a missing value; subsequently, the LFQ intensity was imputed. If samples with missing values exceeded 70% of all samples within a group, the LFQ intensity values were imputed to be 0. If samples with missing values did not exceed 70% of all samples within a group, KNN imputation was performed. The scikit-learn (v0.23.2) Python package was invoked, using the KNNImputer function, which was applied with the neighbor number (k) set to the sample size for the group. Locally developed Python code was used to summarize the results.

Log transformation (base 2) was performed for imputed LFQ intensity values using locally developed Python code. Data were visualized using density distributions and boxplots, which were generated with the matplotlib Python package (version 3.7.5). Hierarchical clustering analysis was performed for all proteins using the pheatmap (version 1.0.12) R package. ggplot2 (v3.5.1) R package was used to generate clustering plots. PCA was performed using the gmodels (version 2.19.1) R package. ggplot2 (v3.5.1) and scatterplot3d (v0.3.44) R packages were used to generate 2D and 3D PCA plots, respectively.

Differential expression analysis was performed using the limma (version 3.62.1) R package. The volcano plot was generated using ggplot2 (v3.5.1) R packages. Heatmaps of differentially expressed proteins were generated using the pheatmap (version 1.0.12) R package. GSEA was performed using the fgsea (version 1.32.2) and clusterProfiler (version 4.14.4) packages. Adjusted P value < 0.05 was adopted in selecting significant enrichment pathways. Pathway enrichment analysis was carried out to identify enriched GO terms among up-/down-regulated proteins, using DAVID (v2023q4) web tools (https://davidbioinformatics.nih.gov/). FDR cut-off of 0.05 was adopted in selecting significant pathways. ggplot2 (v3.5.1) R package was used to generate bubble plots. Gene set enrichment analysis (GSEA) using Gene Ontology (GO) terms, including Biological Process (BP), Cellular Component (CC), and Molecular Function (MF), as well as using the Reactome dataset, for the six pair-wise comparisons between the four groups of samples. Four sets of GSEA were conducted for each of the 6 pair-wise comparisons, for a total of 24 sets of analysis attempted.

#### Microarray data analysis

Each chip had serially diluted anti-IgM and IgM as positive controls to monitor the experimental process, and PBS was used as a negative control. For the obtained chip image, LuxScan 3.0 software was used to read the original data. Statistical tests were then carried out on the chip background, the signal intensity of the positive and negative control sites. The results showed that the Ig control signal value was higher, and the uniformity was good in different samples. In addition, the value of PBS anti-Ig control and background signal value were all low, which met the quality control requirements. The chip of the test sample was scanned with LuxScan 10K-B scanner. Autoantigen microarray chip/ Pathogen-associated antigen microarray chip had 256 points in total. After removal of the 8 anti-IgM and 8 Ig control points, 240 data points were finally obtained. Each protein on the chip was present as two technical repetitions, representing 120 autoantigens. The chip is read by LuxScan 3.0 software to obtain the original data, including foreground signal (F Median), background signal (B Median) and so on. Foreground Median, Background Median columns were extracted from the .LSR file. The fluorescence intensity value of each site was calculated by the formula: Net Fluorescence Intensity (NFI) Value = (Foreground Median - Background Median); SNR = (Foreground Median - Background Median) / SD(Background). SNR was used to filter unreasonable data. The net fluorescence value was set as SNR <0.05 and SNR to 0.001. The net fluorescence value was calculated after subtracting the blank control. The NFI and SNR of the following unreasonable situations were set to 0.001. NFI<20 and SNR>5; SNR<0.05 and NFI>20; NFI<0.05. RLM Normalization was used to normalize the NFI and calculate the effect values of different blocks and slides. For the microarray data clustering and multivariate analysis, first, a log transformation was carried out, and then ‘pheatmap’ was used using the R package with Ward.D2 clustering to make the cluster analysis.

#### Machine learning and Multivariate analysis of autoantigen IgM microarray data

Multivariate analysis of patient clusters based on distance metrics derived from IgM antibody levels against for a panel of autoantigens. Columns with zero variation (constant values) were removed. Log-transformed scaled data showing relative differences between patients was used because the data in its raw form had different orders of magnitude, making analysis and comparison difficult. The analysis was performed using the R package “pheatmap” using Ward.D2 clustering method and Euclidean distances.

A Random Forest classifier using the R package randomForest was fit, predicting ME/CFS as an outcome, on two types of data - raw data and log-scaled data. The purpose of this fit was a rapid screen for variables for further analysis. The Variable Importance Plot for the fits was plotted. The top 10 candidates from the Log Variable Importance Plots were selected for Principal Component Analysis (PCA), a statistical methodology for dimension reduction. PCA was performed using the R package stats (prcomp) and the FactoMineR package.

#### Other statistical analysis

All statistical calculations were performed using GraphPad Prism 10.0. Error bars displayed on graphs represent the means ± SD of three or more independent replicates of an experiment. Statistical significance was calculated separately for each experiment and is described within individual figure legends. For image analysis, three or more biological replicates per sample condition were used to generate the represented data. The results were considered significant at P ≤ 0.05.

## Data Availability

The mass spectrometry proteomics data for the serum immunoglobulin proteome study have been deposited in the ProteomeXchange Consortium via the PRIDE partner repository with the dataset identifier PXD065439.
Any additional information required to reanalyze the data reported in this paper is available from the lead contact upon request.

## Acknowledgement

We thank the Core Unit for Confocal Microscopy and Flow Cytometry-based Cell Sorting at the IZKF Würzburg for their support of this study. We thank Vera Kozjak-Pavlovic (Biocenter, University of Würzburg) for providing a custom macro for mitochondrial analysis. We thank AccuraScience, USA, for their support in mass spectrometry data analysis and Dr. Archana Prusty for her help in setting up the in-house IgG purification pipeline.

## Author Contribution

BKP conceived the idea, developed, and supervised the project, and wrote the manuscript together with ZL.

ZL conducted all IgG purification assays, mitochondrial analysis, and Seahorse assays.

CH contributed to the development of HUVEC cells carrying mitoGFP.

EEB performed simple Western gel runs and analyzed the data.

GN, LS carried out IgG-induced inflammation assays in PBMCs and did cytokine measurement assays.

SK performed multivariate and other statistical analyses of autoantigen IgM microarray data.

RR, JBS, FP, CS, FS, GR, and ZNK contributed to patient recruitment, sample collection, and patient data management.

AS and SL conducted mass spectrometry studies and performed related data analysis.

RN contributed to the design and supervision of the mitochondrial work and data analysis.

All other authors critically revised the manuscript. All authors approved the submitted version.

## SUPPLEMENTARY FIGURE LEGENDS

**Supplementary Figure S1.**
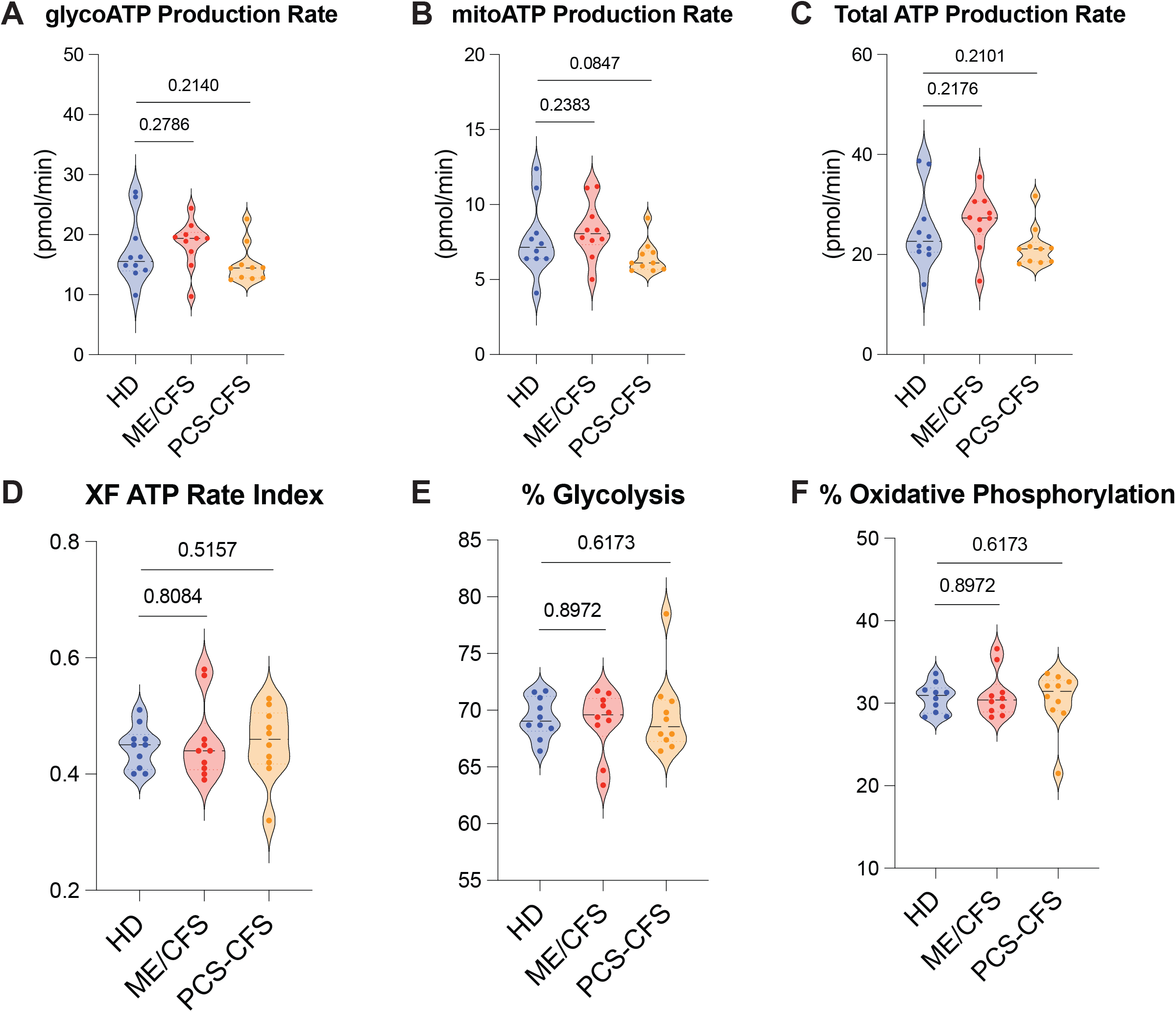
Comparison of ATP generation capacity in HUVEC cells exposed to patient-derived IgG. A-F. Seahorse real-time ATP rate assay in HUVECs exposed to 1⍰μg/mL IgGs from 10 ME/CFS and 10 PCS-CFS patients and 10 controls. A. Glycolytic ATP production rate. HD vs ME/CFS, ns P=0.8109. HD vs PCS-CFS, ns P=0.8109. B. Mitochondrial ATP production rate. HD vs ME/CFS, ns P=0.9264. HD vs PCS-CFS, ns P=0.9860. C. Total ATP production rate. HD vs ME/CFS, ns P=0.7806. HD vs PCS-CFS, ns P=0.7959. D. XF ATP rate index. HD vs ME/CFS, ns P=0.8676. HD vs PCS-CFS, ns P=0.9888. E. ATP from glycolysis (%). HD vs ME/CFS, ns P=0.8955. HD vs PCS-CFS, ns P=0.9263. F. ATP from oxidative phosphorylation (%). HD vs ME/CFS, ns P=0.8955. HD vs PCS-CFS, ns P=0.9263

**Supplementary Figure S2.**
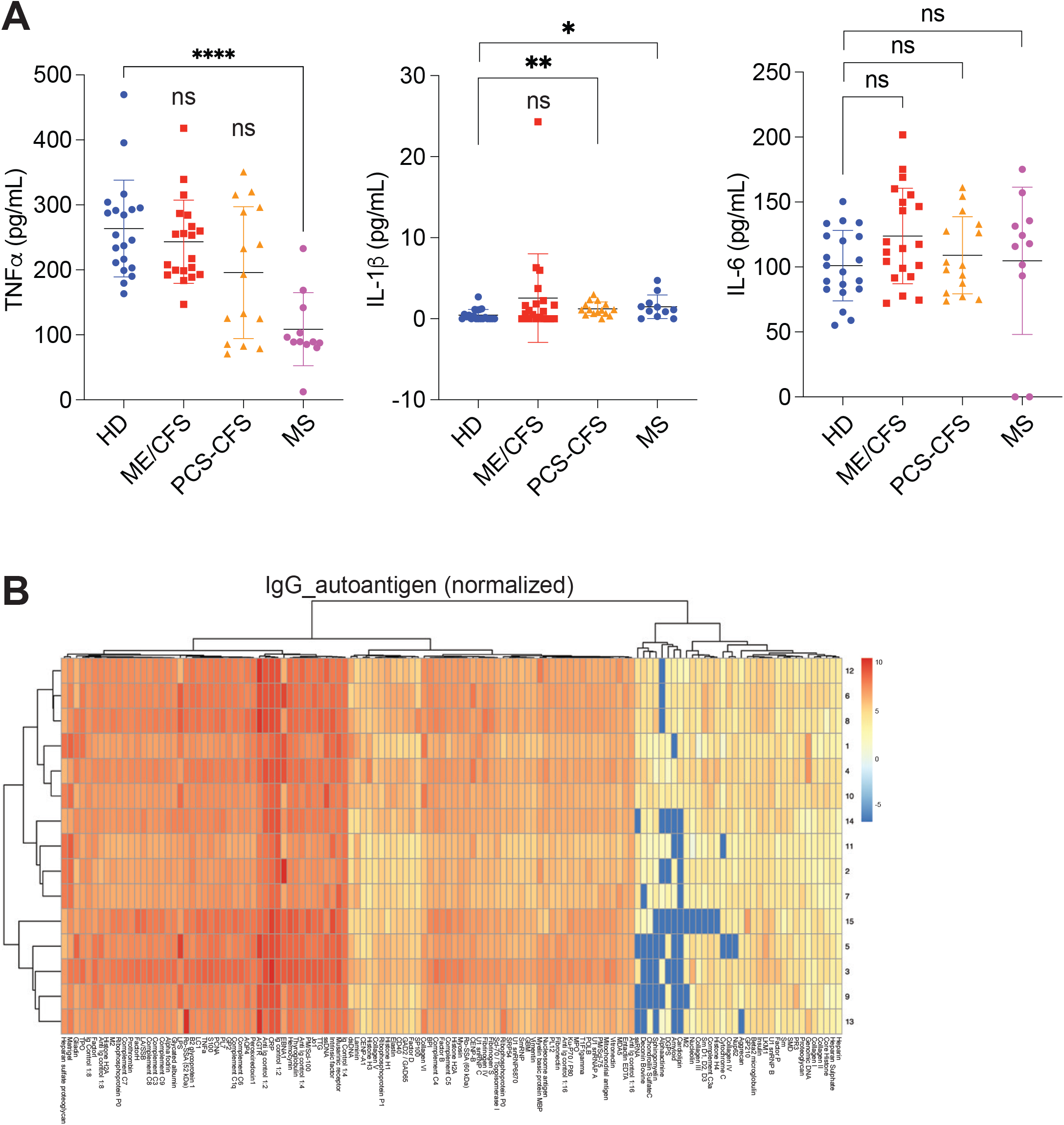
IgG induced inflammatory cytokines and autoantigens. A. ELISA-based validation of secreted cytokine levels of TNF-α, IL-1β, and IL-6 from PBMCs after 48 hours of exposure of PBMCs to IgG from ME/CFS (n=20), PCS-CFS (n=15), MS (n=11), and healthy controls (n=20). (TNF-α) HD vs ME/CFS, ns P=0.3547. HD vs PCS-CFS, ns P=1066. HD vs MS, ****P<0.0001. (IL-1b) HD vs ME/CFS, ns P=0.0887. HD vs PCS-CFS, **P=0.0030. HD vs MS, *P=0.0182. (IL-6) HD vs ME/CFS, ns P=0.0675. HD vs PCS-CFS, ns P=0.5869. HD vs MS, ns P=0.2931. B. Multivariate analysis of clusters based on distance metrics derived from IgG antibody levels for a panel of autoantigens. Log-transformed scaled data showing relative differences between different variables in both healthy controls and patients.

---

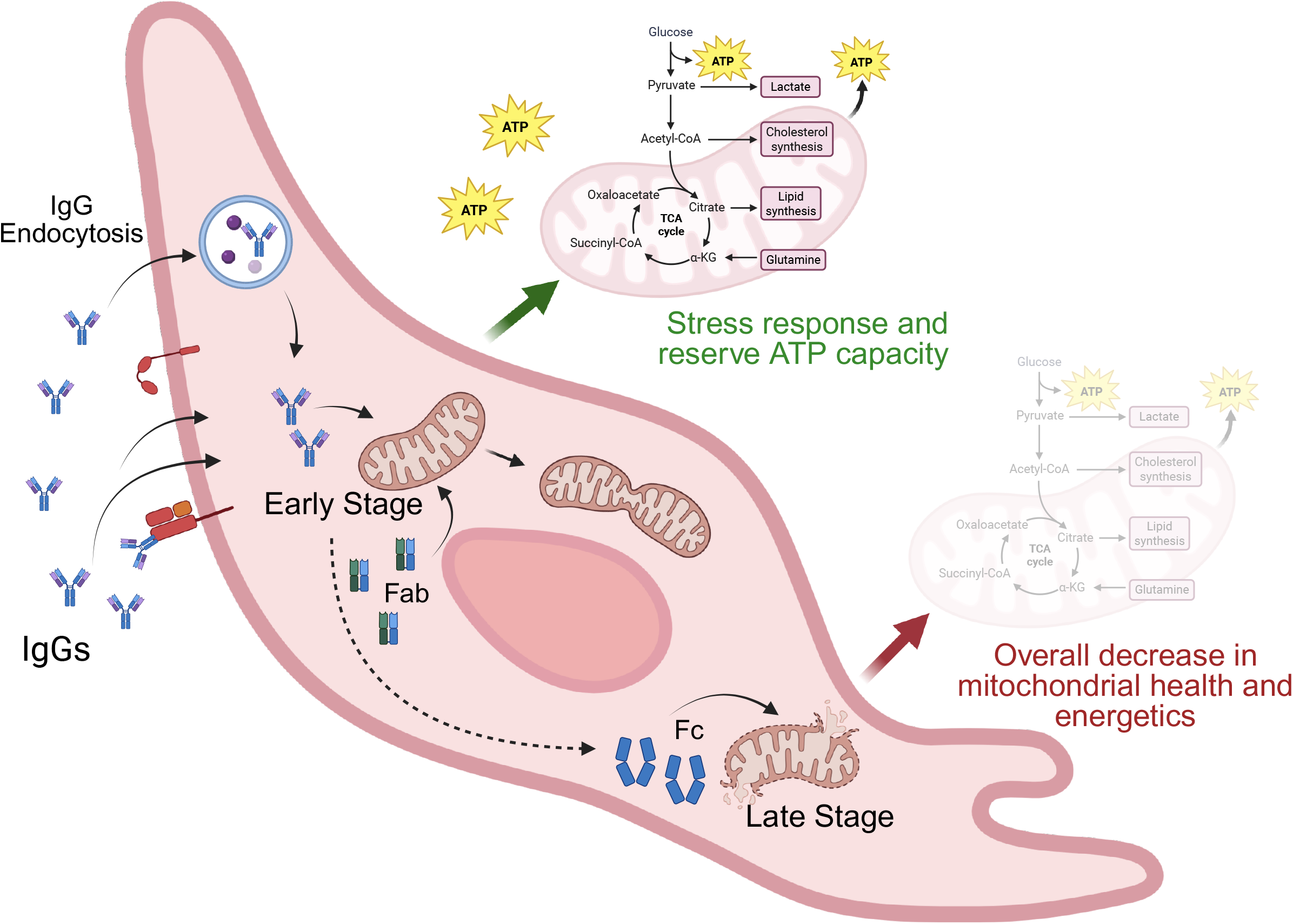

