## Supplementary Tables 1 and 2 for "ME/CFS and PASC Patient-Derived Immunoglobulin Complexes Disrupt Mitochondrial Function and Alter Inflammatory Marker Secretion"

**Supplementary Table 1:** Patient demographics.

The whole study cohort:

| Demographics | | HD (n = 40) | ME/CFS (n = 41) | PCS-CFS (n = 16) | MS (n = 20) |
|---|---|---|---|---|---|
| Gender | Female | 27 | 28 | 11 | 16 |
| | Male | 13 | 13 | 5 | 4 |
| Age | Mean ± SD | 40.93 ± 12.17 | 39.85 ± 8.50 | 40.75 ± 10.34 | 38.70 ± 9.97 |
| | Median (min-max) | 39 (12-65) | 39 (22-59) | 41.5 (21-58) | 40 (20-54) |
| Bell Score | 10-20 | 0 | 14 | 5 | 0 |
| | 30-40 | 0 | 21 | 8 | 0 |
| | 45-60 | 0 | 6 | 1 | 0 |
| | na | 40 | 0 | 2 | 20 |

Mass spectrometry study:

| Demographics | | HD (n = 39) | ME/CFS (n = 40) | PCS-CFS (n = 16) | MS (n = 11) |
|---|---|---|---|---|---|
| Gender | Female | 26 | 28 | 11 | 9 |
| | Male | 13 | 12 | 5 | 2 |
| Age | Mean ± SD | 41.13 ± 12.26 | 39.88 ± 8.61 | 40.75 ± 10.34 | 36.27 ± 9.33 |
| | Median (min-max) | 39 (12-65) | 39 (22-59) | 41.5 (21-58) | 38 (20-45) |
| Bell Score | 10-20 | 0 | 13 | 5 | 0 |
| | 30-40 | 0 | 21 | 8 | 0 |
| | 45-60 | 0 | 6 | 1 | 0 |
| | na | 39 | 0 | 2 | 11 |

Mitochondrial study:

| Demographics | | HD (n = 39) | ME/CFS (n = 41) | PCS-CFS (n = 15) | MS (n = 20) |
|---|---|---|---|---|---|
| Gender | Female | 27 | 28 | 16 | 16 |
| | Male | 12 | 13 | 4 | 4 |
| Age | Mean ± SD | 41.31 ± 12.08 | 39.85 ± 8.50 | 38.70 ± 9.97 | 38.70 ± 9.97 |
| | Median (min-max) | 39 (12-65) | 39 (22-59) | 40 (20-54) | 40 (20-54) |
| Bell Score | 10-20 | 0 | 14 | 0 | 0 |
| | 30-40 | 0 | 21 | 0 | 0 |
| | 45-60 | 0 | 6 | 0 | 0 |
| | na | 39 | 0 | 20 | 20 |

**Supplementary Table 2:** List of antigens used in the microarray study.

| Supplementary Table 2 |
|---|
| **Autoantigens** |
| Aggrecan |
| AGTR |
| Alpha Fodrin |
| alpha-actinine |
| Amyloid |
| AQP4 |
| B2 glycoprotein 1 |
| B2-microglobulin |
| BPI |
| Cardolipin |
| CD40 |
| CENP-A |
| CENP-B |
| Chondroitin Sulfate C |
| Collagen I |
| Collagen II |
| Collagen III |
| Collagen IV |
| Collagen V |
| Collagen VI |
| complement C1q |
| complement C3 |
| complement C3a |
| complement C4 |
| complement C5 |
| complement C6 |
| complement C7 |
| complement C8 |
| complement C9 |
| Core Histone |
| CRP |
| Cytochrome C |
| Decorin-bovine |
| DGPS |

| |
|---|
| DNA Polymerase beta (POLB) |
| dsDNA |
| EBNA1 |
| Elastin |
| Entaktin EDTA |
| Factor B |
| Factor D |
| Factor H |
| Factor I |
| Factor P |
| Fibrinogen IV |
| Fibrinogen S |
| Fibronectin |
| GAD2/GAD65 |
| GBM |
| Genomic DNA |
| Gliadin |
| Glycated Albumin |
| GP2 |
| GP210 |
| Hemocyanin |
| Heparan sulfate proteoglycan |
| Heparan Sulphate |
| Heparin |
| Histone H1 |
| Histone H2A |
| Histone H2B |
| Histone H3 |
| Histone H4 |
| Insulin |
| Intrinsic Factor |
| Jo-1 |
| KU (P70/P80) |
| La/SSB |
| Laminin |
| LC1 |
| LKM1 |
| LPS |

| |
|---|
| M2 |
| Matrigel |
| MDA5 |
| Mi-2 |
| Mitochondrial antigen |
| MPO |
| Muscarinic receptor |
| Myelin basic protein (MBP) |
| Myosin |
| Nucleolin |
| Nucleosome antigen |
| Nup 62 |
| PCNA |
| Peroxiredoxin 1 |
| PL-7 |
| PL-12 |
| PM/Scl 100 |
| PM/Scl-75 |
| PR3 |
| Proteoglycan |
| Prothrombin protein |
| Ribo Phosphoprotein P0 |
| Ribo Phosphoprotein P1 |
| Ribo Phosphoprotein P2 |
| Ro/SSA (52 Kda) |
| Ro/SSA (60 Kda) |
| S100 |
| Scl-70/Topoisomerase I |
| Sm |
| Sm/RNP |
| SmD |
| SmD1,D2,D3 |
| SP100 |
| Sphingomyelin |
| SRP54 |
| ssDNA |
| ssRNA |
| T1F1 gama |
| Thyroglobulin |

| |
|---|
| TNF-a |
| TPO |
| TTG |
| U1-snRNP 68/70 |
| U1-snRNP A |
| U1-snRNP B/B U1-snRNP B/B |
| U1-snRNP C |
| Vimentin |
| Vitronectin |
